# Common genetic variants near *SLC2A2* and glycemic response to glimepiride in the GRADE comparative effectiveness clinical trial

**DOI:** 10.1101/2025.10.07.25336827

**Authors:** Josephine H. Li, Lukasz Szczerbinski, Mark Tripputi, Haiyin Liu, Stella Nam, Endrina Mujica, Anastasia Emmanouilidou, Thinley Yidzin Wangden, Anna Citko-Rojewska, Paulina Konopka, Marcin Czajkowski, Alicia Huerta-Chagoya, Maheak Vora, Aaron Leong, James B. Meigs, Maggie Y. Ng, Ruth J.F. Loos, Marie Pigeyre, Hertzel C. Gerstein, Filipe A. Moura, Yi-Pin Lai, Deepak L. Bhatt, Nicholas A. Marston, Christian T. Ruff, Marc S. Sabatine, Adam Y. Dawed, Ewan R. Pearson, Leslie S. Satin, Marcel den Hoed, Adam Kretowski, Steven E. Kahn, Naji Younes, Josep M. Mercader, Jose C. Florez, the GRADE Research Group

**Author notes:** <u>Corresponding author:</u> Jose C. Florez, MD, PhD; 185 Cambridge St. Boston, MA 02114. Contributed equally to this work (Josephine H. Li, Lukasz Szczerbinski). Jointly directed this work (Josep M. Mercader, Jose C. Florez). GRADE Research Group Listing provided as Supplemental Item 1.

## Abstract

Optimizing second-line therapy for type 2 diabetes is challenging due to interindividual variability in response. We conducted a pharmacogenomic genome-wide association study (GWAS) in the Glycemia Reduction Approaches in Type 2 Diabetes: A Comparative Effectiveness (GRADE) Study to identify genetic predictors of glycemic response to insulin glargine, glimepiride, liraglutide, and sitagliptin, when added to metformin in a diverse population. We identified 21 genome-wide significant loci associated with treatment response. rs1905505, a non-coding variant near *SLC2A2*, the gene encoding the glucose transporter GLUT2, was enriched in Africans/African Americans and conferred a 36% increased risk of treatment failure on glimepiride (*p*=4.83×10). Carriers had impaired β-cell function, evidenced by a lower C-peptide index during OGTT, and diminished glucose-lowering response to an acute sulfonylurea challenge. Genetic manipulation in zebrafish confirmed that *slc2a2* disruption attenuates the glucose-lowering effect of glimepiride. In conclusion, genetic variation influences glycemic response to medications, with *SLC2A2* emerging as a key determinant of sulfonylurea response.

Clinical Trial registration number: NCT01794143

## Background

A major challenge in treating type 2 diabetes (T2D) is selecting the optimal medication to achieve and maintain glycemic control. Over 20% of patients initially prescribed metformin might require escalation of therapy at 5 years.^1^ The Glycemia Reduction Approaches in Diabetes: A Comparative Effectiveness (GRADE) Study is a multicenter clinical trial that compared the effectiveness of four classes of commonly-prescribed glucose-lowering medications (basal insulin, sulfonylurea, glucagon-like peptide-1 receptor agonist [GLP-1RA], and dipeptidyl peptidase-4 inhibitor [DPP4-i]) as second-line therapy.^2^ The basal insulin glargine and GLP-1RA liraglutide were most effective in maintaining glycated hemoglobin (HbA1c) levels within recommended ranges, while sitagliptin was least effective;^3^ however, the glycemic response to each medication varied between individuals.

The heritability of glycemic response to metformin and sulfonylureas has been estimated to be 34–37%,^4,5^ supporting the influence of genetic variation on the response to glucose-lowering medications. Genome-wide association studies (GWAS) for response to metformin, sulfonylureas, and GLP-1RAs have revealed genetic effects with biological significance^5–7^ and the potential to influence clinical practice. Yet, treatment selection does not consider genetic information.

In this pharmacogenomic discovery study, we searched for genetic variants associated with glycemic response to each of the GRADE-assigned medications by conducting a GWAS of treatment response, defined by the time to reach primary (HbA1c≥7%) and secondary (HbA1c>7.5%) metabolic failure (Figure 1). We then sought to replicate each medication-specific set of genome-wide significant variants in independent cohorts, including the Outcome Reduction with an Initial Glargine Intervention (ORIGIN) trial,^8^ the Saxagliptin Assessment of Vascular Outcomes Recorded in Patients with Diabetes Mellitus (SAVOR)–Thrombolysis in Myocardial Infarction (TIMI) 53 trial,^9^ and published GWAS meta-analyses of sulfonylurea and GLP-1RA responses.^5,7^ Mechanistic follow-up analyses investigated the effect of the top variants and *SLC2A2* gene function through 1) serial oral glucose tolerance tests (OGTTs) in GRADE, 2) an independent acute challenge with glipizide, a sulfonylurea closely related to glimepiride, in the Study to Understand the Genetics of the Acute Response to Metformin and Glipizide in Humans (SUGAR-MGH),^10^ and 3) analysis of glimepiride effects in zebrafish CRISPR/Cas9 *slc2a2* mutant models.

**Figure 1.**
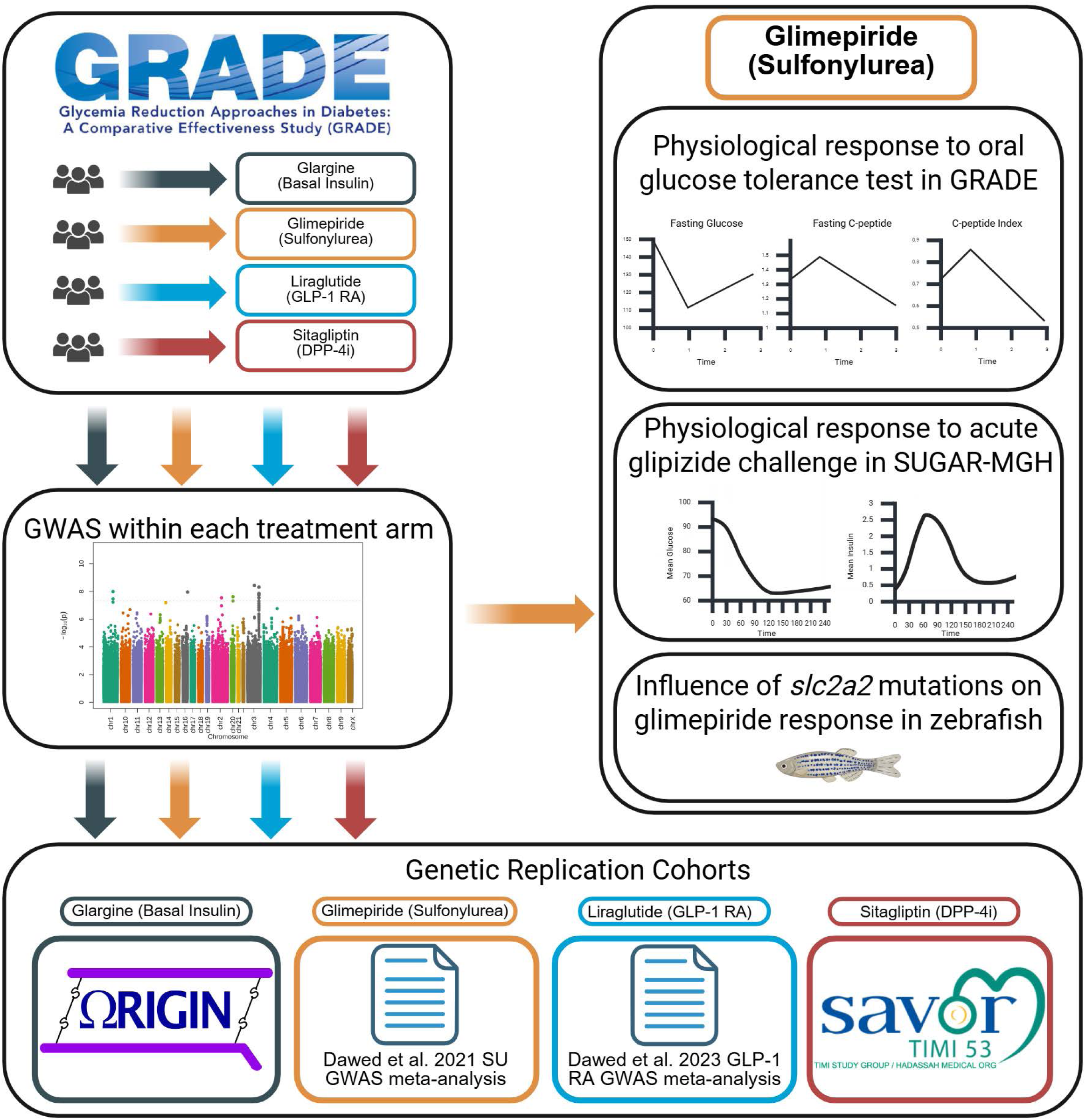
Overview of the analysis strategy.

## RESULTS

### Baseline characteristics

Participants included in the genetic analysis (n=4,572, 90.6% of the full cohort, 68.7% White, 18.4% Black or African American, 12.9% other self-reported race, and 18.9% Hispanic) were mostly middle-aged (57.5±9.9 years) and male (64.7%). Compared to other treatment groups, participants randomized to glargine had higher fasting insulin at baseline (*p*=0.01, Table S1). Other baseline characteristics did not differ by treatment group. Compared to those who did not consent to genetic analyses, participants included in genetic analyses were older, included a greater proportion of men, and were more likely to self-report as White (*p*<0.001, Table S2). Across the 36 clinical centers, there were significant differences in the proportion of participants who did not consent or had insufficient DNA quality (*p*<0.001).

### Loci associated with GRADE outcomes across treatment groups

Our time-to-event GWAS for primary (HbA1c≥7%) and secondary (HbA1c>7.5%) outcomes across the four treatment groups identified three loci significantly (*p*<5×10^-8^) associated with response to sitagliptin, six for glargine, four for liraglutide, and eight for glimepiride (Table 1). For sitagliptin, rs11409687 near *ADAMTS1* was associated with a higher risk of reaching the secondary outcome (hazard ratio [HR] 2.40, 95% CI 1.76-3.27, *p*=3.17×10^-8^, Figure S2A-B). For liraglutide, rs115566325 near *ZNF217* was associated with a higher risk of reaching both primary (HR 2.15, 95% CI 1.64-2.84, *p*=4.64×10^-8^) and secondary (HR 2.66, 95% CI 1.94-3.66, *p*=1.66×10^-9^, Figure S2C-D) outcomes. For glargine, rs10423332 within *TMEM91* was associated with a higher risk of reaching the secondary outcome (HR 3.50, 95% CI 2.27-5.41, *p*=1.68×10^-8^, Figure S2E-F). In the glimepiride group, we identified significant signals at rs11925227 and rs1905505 on chromosome 3 near *SLC2A2*, which encodes the glucose transporter 2 (GLUT2, Figure 2A). Both were associated with an increased risk of treatment failure, with the rs11925227 A allele conferring a HR of 1.45 (95% CI: 1.28–1.65, *p*=4.87×10^-9^), and the rs1905505 A allele with a HR of 1.36 (95% CI: 1.22–1.52, *p*=4.83×10^-8^; Figure 2B).

**Figure 2.**
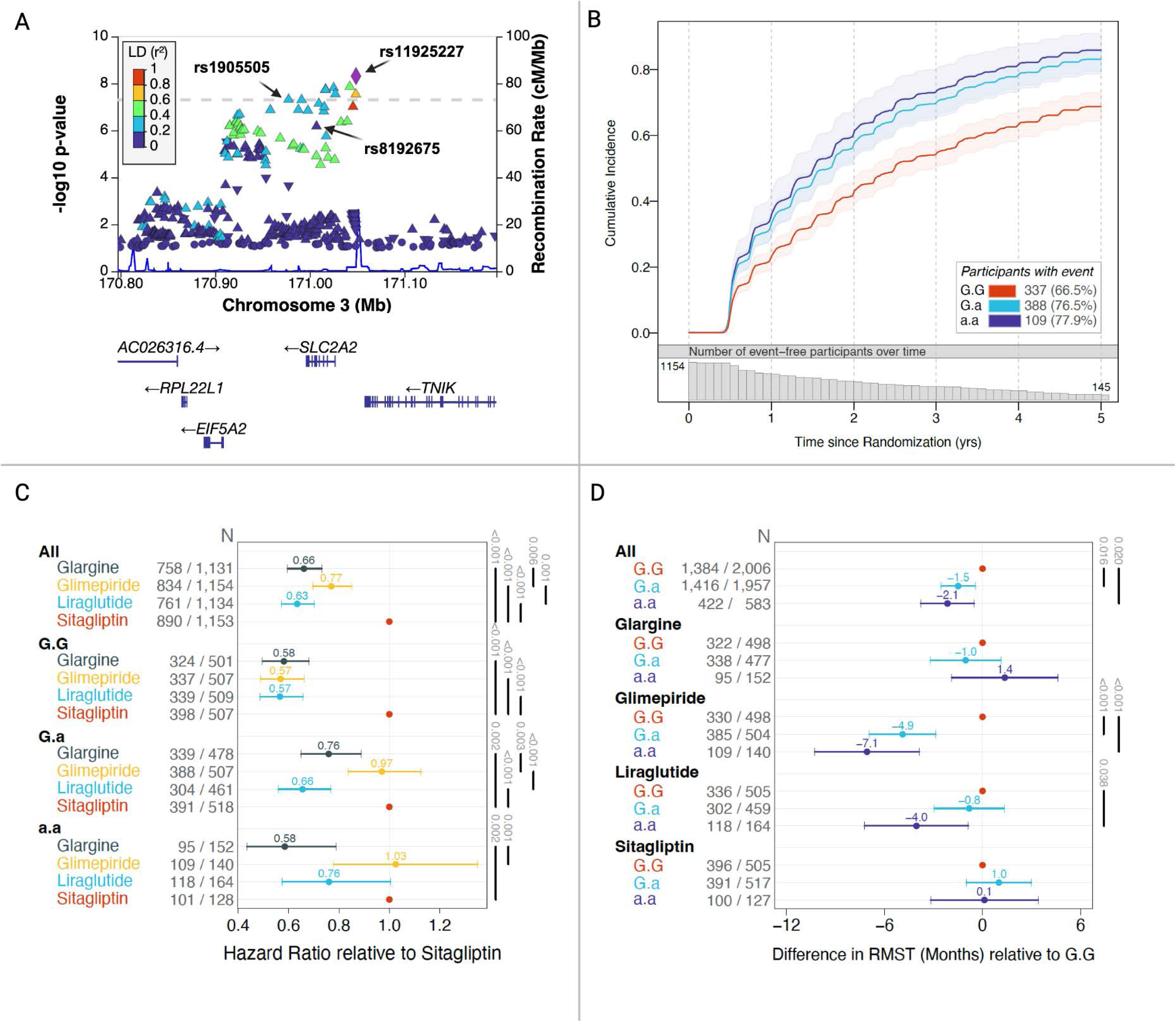
A) Regional association plot of rs1905505, rs11925227, and rs8192675 for primary metabolic outcome in the glimepiride group. B) Cumulative incidence plots for the primary metabolic outcome in the glimepiride group by rs1905505 genotype. C) Hazard ratios (HRs) of treatment failure, as measured by reaching the primary metabolic outcome, stratified first by rs1905505 genotype, followed by treatment group. Sitagliptin is the reference group with a HR of 1.0. D) Restricted mean survival time (RMST) of treatment failure, as measured by months to reaching the primary metabolic outcome, stratified first by treatment group, followed by rs1905505 genotype. Differences in RMST are reported relative to non-carriers of the A effect allele.

**Table 1.**
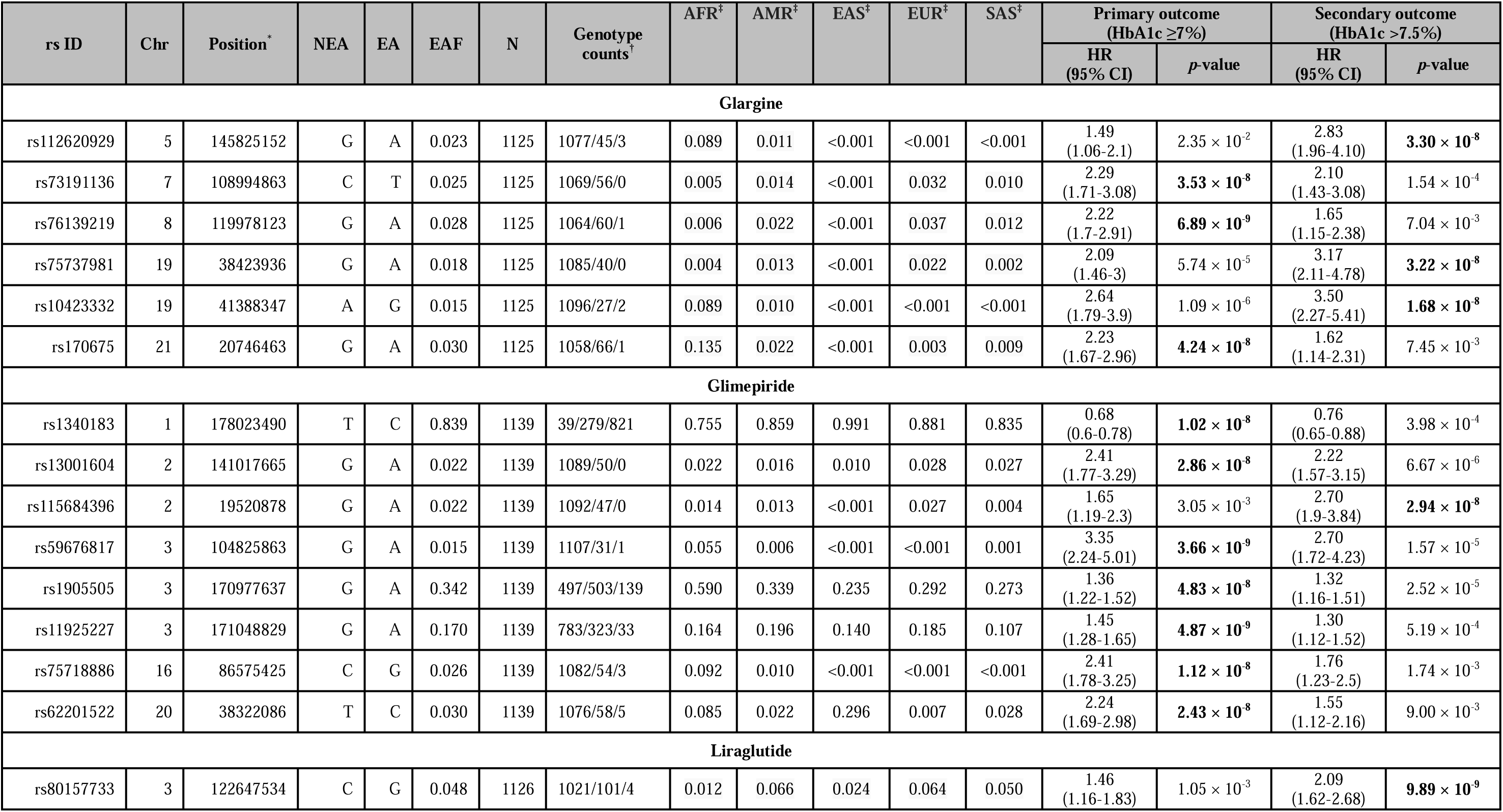
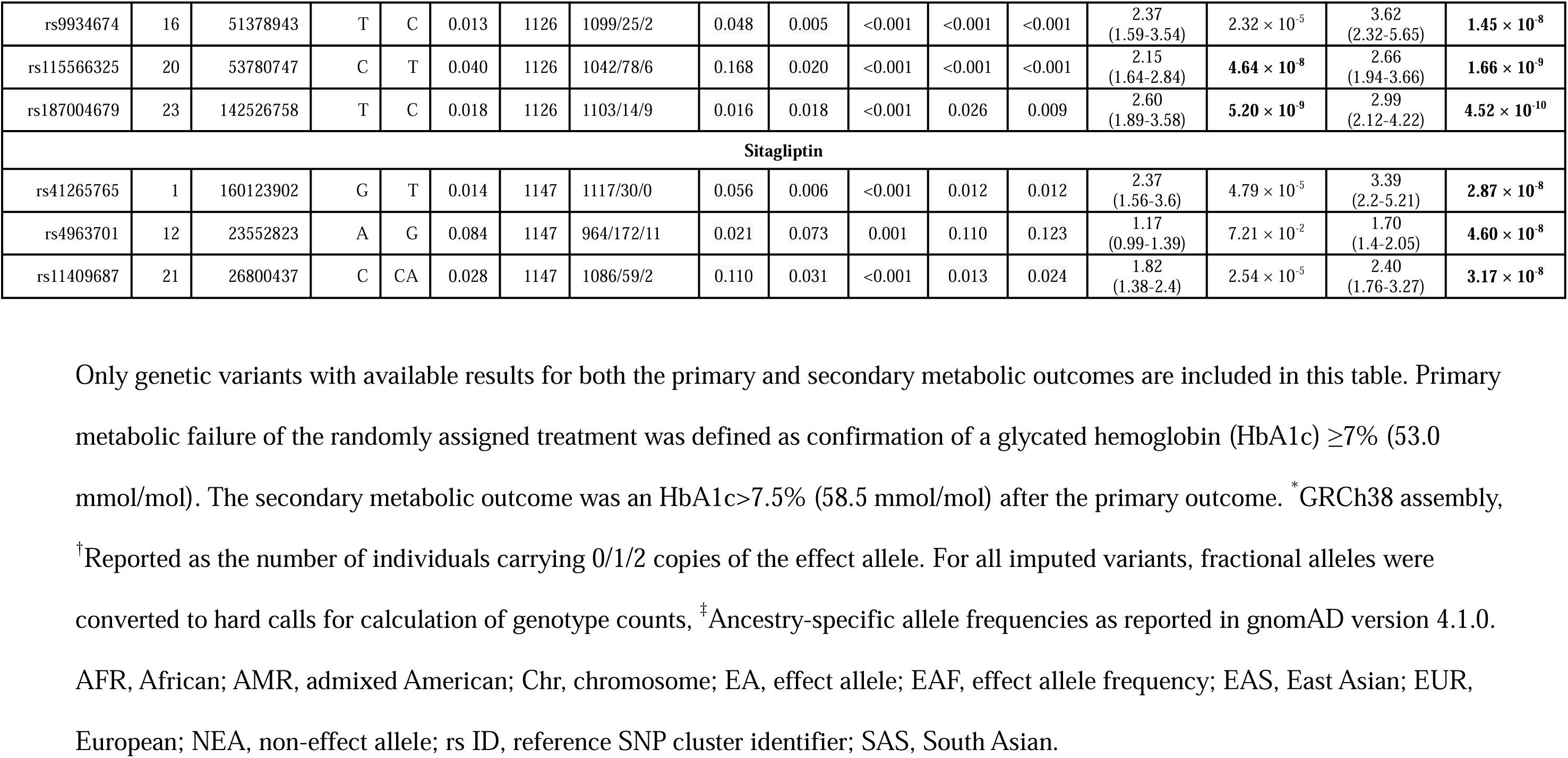
Genome-wide significant findings (*p*<5×10^-8^) for the primary and secondary metabolic outcomes by treatment group.

### Independent replication

Of the 21 variants from the GRADE GWAS, only 15 were present in the independent clinical trial replication datasets used in this report. In a replication analysis we conducted in a published sulfonylurea-response GWAS,^5^ we observed a 0.04% less HbA1c reduction per A allele in response to a sulfonylurea (*p*=0.04). While the association did not persist after multiple testing, it was consistent with our findings that the same A allele increased risk of the primary outcome in GRADE (Table S3). The only liraglutide variant available for look-up did not reach nominal significance in the replication cohort (Table S4). Of six variants associated with glargine response in GRADE, four were observed in ORIGIN: two reached nominal significance but in the opposite direction (Table S5). The three variants associated with sitagliptin response in GRADE did not replicate in the GWAS of glycemic response to the DPP4-i saxagliptin in SAVOR-TIMI 53 (Table S6).

### Variants near *SLC2A2* are associated with glimepiride failure

We observed the effect allele frequency (EAF) for rs1905505 was higher in African/African American (AFR) compared to European genetic ancestries (EUR) (AFR_EAF_=0.59, EUR_EAF_=0.29), while the EAF for rs11925227 was similar (AFR_EAF_=0.16, EUR_EAF_=0.19). Linkage disequilibrium (LD) between rs11925227 and rs1905505 differed by ancestry (AFR, r²=0.09; EUR, r²=0.55; Figure S3). To estimate the independent contributions of these variants, we performed conditional analyses. When conditioning on rs11925227, rs1905505 remained significantly associated with treatment failure in the overall population and self-reported White participants (Table S7). When conditioning on rs1905505, rs11925227 retained significance in the overall population (HR 1.27, 95% CI 1.09–1.49, *p*=2×10^-3^) but not among Whites (HR 1.13, 95% CI 0.92–1.39, *p*=0.24), suggesting rs1905505 is more likely to be the causal variant.

Non-risk allele carriers at rs1905505 (GG genotype) showed increased responsiveness (42-43% reduced risk of treatment failure) to glargine, liraglutide, and glimepiride compared to sitagliptin, the least effective medication in GRADE (Figure 2C).^3^ However, carriers of the risk allele (GA and AA) showed reduced effectiveness to glimepiride, similar to sitagliptin, an effect not seen for liraglutide and glargine.

To determine the time to treatment failure by genotype, we compared the restricted mean survival time (RMST). In the glimepiride group, the RMST (in years) was 2.5 (95% CI 2.3-2.6) for GG, 2.0 (95% CI 1.9-2.2) for AG, and 1.9 (95% CI 1.6-2.1) for AA carriers. Compared to GG carriers, AG and AA carriers reached the primary outcome 4.9 months and 7.1 months earlier, respectively (*p*<0.001, Figure 2D). Notably, in the liraglutide group, rs1905505 had a similar effect, with a 34% increased risk of treatment failure in AA carriers compared to GG carriers (*p*=0.046, Figure S4A).

We next characterized variant carriers for which OGTT was available at baseline and at 1 and 3 years after randomization. At baseline, fasting glucose and fasting C-peptide were similar by rs1905505 genotype in the glimepiride group (Figure 3A-B). However, A-allele carriers had a lower C-peptide index (CPI, *p*<0.001), indicating poorer β-cell function after a glucose challenge before glimepiride treatment (Figure 3C). This difference in CPI by genotype was exacerbated at years 1 and 3 after glimepiride treatment, with greater separation between genotype groups, showing lower fasting C-peptide and less reduction of fasting glucose levels in A-allele carriers. Similar differences in CPI by genotype were observed in the liraglutide group (Figure S5). The as-treated analyses were consistent with the intention-to-treat analyses (Figure S6).

**Figure 3.**
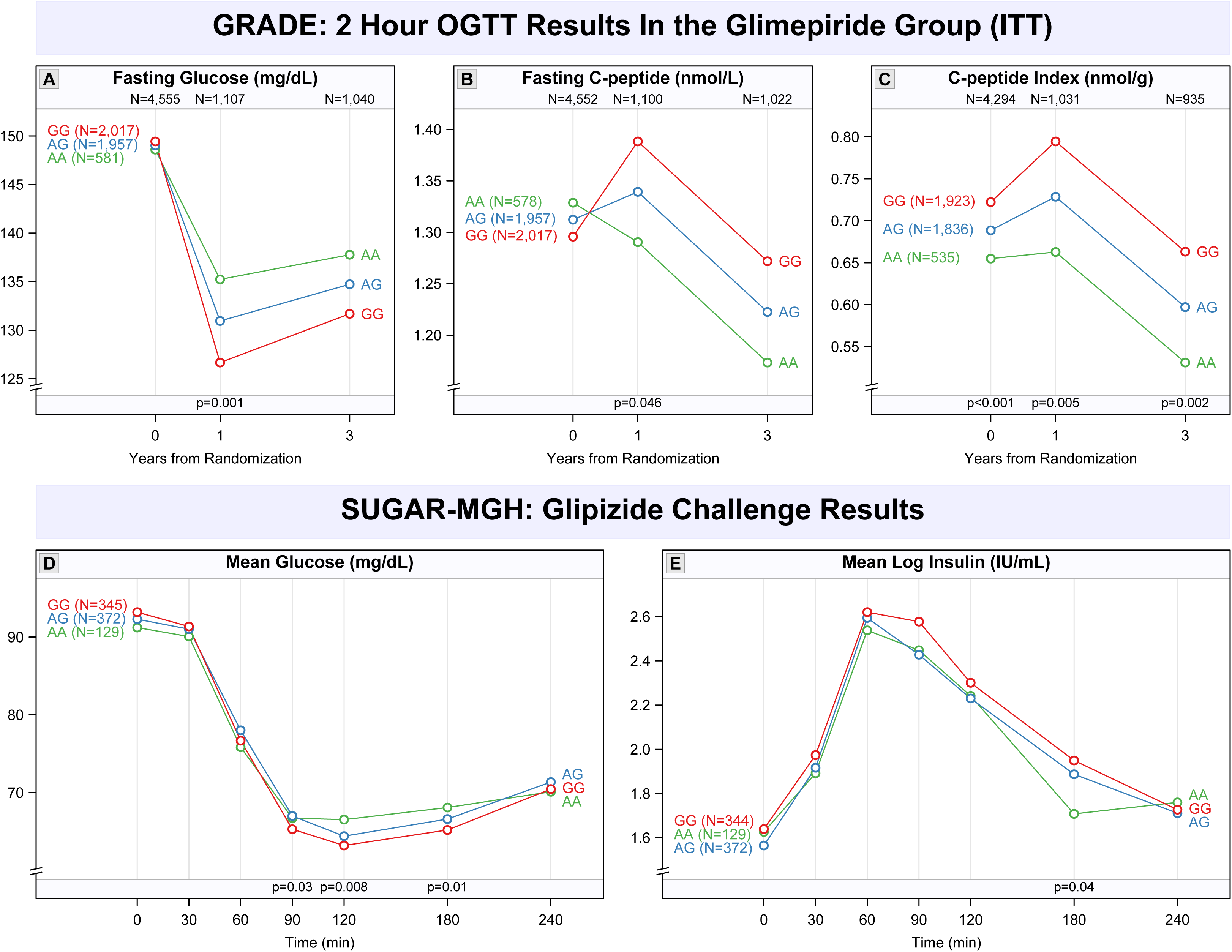
A) Fasting glucose, B) Fasting C-peptide, and C) C-peptide index (CPI) during the OGTT performed at years 0, 1, and 3 from randomization in GRADE by rs1905505 genotype. The reported N’s for AA, GG, and AA represent the distribution of rs1905505 genotype at baseline (year 0) prior to treatment assignment. OGTT data at years 1 and 3 are displayed for the glimepiride group based on intention-to-treat (ITT) analysis. *P*-values compare race-adjusted means of fasting glucose, fasting C-peptide, and CPI by rs1905505 genotype using an additive genetic model. D) Change in glucose (mg/L) and E) log-insulin (IU/mL) by rs19005505 genotype following glipizide administration in SUGAR-MGH. *P*-values reported are for the time × variant interaction term in the mixed-effects model. Error bars represent standard errors.

### rs1905505 is associated with worse response to an acute glipizide challenge

We assessed the association of rs1905505 with the glucose and insulin response to a single 5 mg dose of glipizide, a sulfonylurea, in SUGAR-MGH.^10,11^ On average, A-allele carriers had significantly higher glucose levels at 90, 120, and 180 minutes (Figure 3D) and lower insulin levels at 180 minutes (Figure 3E) after glipizide, indicating a less robust response to an acute sulfonylurea challenge.

### CRISPR/Cas9-induced mutations in *slc2a2* attenuate glycemic response to glimepiride

The rs1905505 A allele was robustly associated with lower *SLC2A2* expression in liver (*p*=3×10^-12^)^12^ and pancreatic islets (*p*=0.01) in one study^13^, while not confirmed in a larger pancreatic islet eQTL dataset,^14^ suggesting that *SLC2A2* is a likely effector gene. To examine whether *SLC2A2* influences differential glimepiride response, we performed a combined gene perturbation and glimepiride treatment experiment in zebrafish larvae. Results from 351 overfed zebrafish larvae showed that CRISPR/Cas9-induced pLOF mutations in *slc2a2* induced a 0.36±0.16 SD units higher random glucose content (*p*=0.03), while 2 days of treating wild-type larvae with 50 μM glimepiride resulted in 0.53±0.17 SD units lower glucose content by day 10 (*p*=0.002). Strikingly, the glucose reduction effect of glimepiride was abolished in *slc2a2* crispants (−0.03±0.13, *p*=0.81, Figure 4), revealing a significant gene×medication interaction (*p*=0.02).

**Figure 4.**
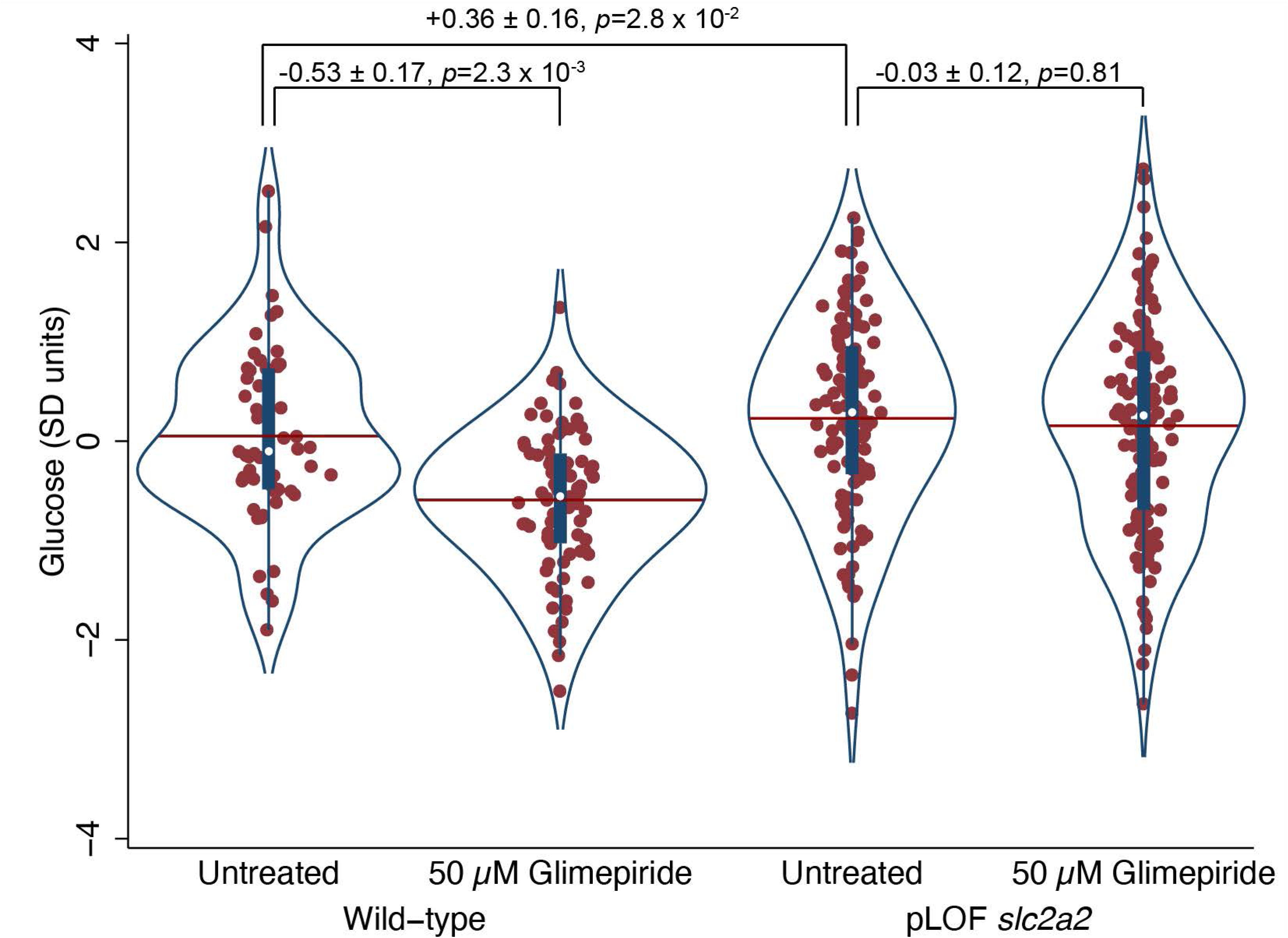
Violin plot showing the main and interaction effects of treatment with 50 µM glimepiride and CRISPR/Cas9-induced pLOF mutations in *slc2a2* on inverse normally transformed random glucose content in 351 overfed, 10-day-old zebrafish larvae. A significant gene×medication interaction was observed (*p*=0.02).

## DISCUSSION

Treatment choices in T2D seldom consider the underlying molecular pathophysiology of the individual patient. Thus, selecting an effective second-line therapy for T2D does not account for potential individual differences in medication response. As genetic factors may partially explain this observed heterogeneity, we searched for genetic predictors of the glycemic response to glargine, glimepiride, liraglutide, and sitagliptin when added to metformin in a diverse T2D population in GRADE. We identified 21 associations between genetic variants and treatment response across the four groups. The lead variant associated with metabolic failure in the glimepiride group (rs1905505), is located 18 kb downstream of *SLC2A2*, which encodes GLUT2, the major glucose transporter in pancreatic β-cells and hepatocytes.^15^ We therefore focused on elucidating this variant’s effects.

Each copy of the rs1905505 A allele conferred a 36% increased risk of glimepiride failure. Moreover, genotype explained some of the heterogeneity in treatment response. When assessed in the full GRADE cohort, glimepiride had lower glycemic efficacy compared to glargine and liraglutide, but its efficacy was comparable to that of glargine and liraglutide in non-carriers of the rs1905505 A allele. On the other hand, in A-allele carriers, glycemic efficacy of glimepiride was indistinguishable from that of sitagliptin, which had the highest overall failure rates in GRADE, suggesting that rs1905505 may have a substantial contribution to the lower glycemic response observed in the glimepiride group.

Multiple lines of evidence further support the association of rs1905505 with glimepiride response. First, we replicated the results in a published sulfonylurea GWAS.^5^ Second, A-allele carriers showed impaired glucose response to an acute sulfonylurea challenge,^11^ consistent with glycemic response to long term treatment in GRADE. While association of rs1905505 with *SLC2A2* expression in pancreatic islets have conflicting results (possibly due to limited power in these datasets),^13,14^ the rs1905505 A allele was robustly associated with lower *SLC2A2* expression in liver,^12^ pointing to this gene as the most likely effector gene for this variant. Thus, we found that CRISPR/Cas9-induced pLOF mutations in *slc2a2* in zebrafish larvae attenuated the decrease in glucose content induced by glimepiride treatment. Finally, rs1905505 is associated with a reduced risk of T2D,^16^ lower random glucose,^17^ and lower HbA1c,^18^ suggesting that the same signals may underlie both the associations with reduced T2D risk and worse response to glimepiride. This suggests that the mechanism driving risk for T2D in this locus may be partially shared by that of sulfonylurea action.

The question remains as to why a genetic variant influencing *SLC2A2* expression impacts the response to sulfonylureas, which are known to promote insulin secretion by inhibiting the ATP-sensitive potassium channel, independently of glucose levels and downstream of GLUT2.^19^ The ATP-sensitive potassium channel-mediated pathway of pancreatic β-cell insulin secretion serves as the initial triggering mechanism for glucose-induced insulin secretion.^20^ However, there is a secondary metabolic amplifying pathway, by which glucose further enhances insulin secretion independent of calcium.^21^ Consequently, GLUT2 plays a role in glucose potentiation, whereby GLUT2 function can amplify glucose-induced insulin secretion by influencing the amount of glucose uptake, thus modulating the efficacy of ATP-sensitive potassium channels closed by sulfonylureas. This is illustrated by our OGTT findings for rs1905505: A-allele carriers, who have lower *SLC2A2* expression leading to impaired glucose transport via GLUT2 and therefore providing less substrate (glucose) for the amplifying pathway, demonstrate a lower CPI, indicative of reduced insulin secretion during the baseline (medication-naïve) OGTT. After treatment with glimepiride, an insulin secretagogue that requires glucose for potentiation of secretion, the effect induced by genetically impaired glucose transport is accentuated, with significantly lower CPI at years 1 and 3 in A-allele carriers compared to non-carriers. These findings were recapitulated to a lesser extent for liraglutide, another insulin secretagogue, in which there was a more modest effect of the rs1905505 variant on treatment failure and a similar pattern seen during the OGTT.

Interestingly, a previous study identified that the C allele of a nearby *SLC2A2* variant, rs8192675, was associated with enhanced glycemic response to metformin.^6^ In GRADE, this same C allele is associated with reduced response to glimepiride when added to metformin monotherapy (HR=1.33, *p*=6.64×10^-7^). rs8192675 is in strong LD with our lead variant rs1905505 in EUR (r²=0.98), whereas LD is lower in AFR (r²=0.58). Our findings suggest that variants in this locus affect glycemic response to glimepiride oppositely compared to the response to metformin observed by *Zhou* et al.^6^ Whether a common mechanism and tissue of action underlies the locus’ effect on response to metformin and glimepiride (when added to metformin) remains to be elucidated.

Our finding of a genetic variant associated with worse glimepiride response may have public health implications globally. While new medication classes like GLP-1RAs and SGLT2 inhibitors are becoming first-line treatments for T2D in developed countries, their high cost limits their use in developing nations, especially in Africa.^22,23^ Since >75% of adults with T2D live in low- or middle-income countries,^24^ cost and ease of administration are primary drivers of medication selection, with metformin and sulfonylureas representing mainstays of therapy.^25^ The lead glimepiride variant is enriched in African/African American ancestry participants, with 83% being either heterozygous or homozygous for the poor-response allele, compared to ∼50% of European ancestry participants. Moreover, homozygotes experienced treatment failure over 7 months earlier compared to non-carriers. Early treatment failure may negatively impact perceived treatment efficacy, making adherence and patient engagement more challenging.^26^ However, we acknowledge that the progression to the primary metabolic outcomes was overall high across variant groups; thus, inclusion of genetic data in medication selection may only yield modest improvements in glycemic outcomes.

Our study had some limitations. Since enrolled participants had T2D for <10 years and a baseline HbA1c of 6.8-8.5% (50.8-69.4 mmol/mol) on ≥1000 mg/day metformin before second-line therapy, it is unclear whether our findings are generalizable to those with long-standing diabetes on more complex regimens. We noted small, albeit significant, differences in baseline characteristics between those included in genetic analyses compared to the remainder of GRADE, such as older age, fewer females, and a higher proportion of self-reported White individuals in those who were genotyped. However, we felt that these differences could have been related to heterogeneity in recruitment practices, as the proportion of participants who consented to genotyping was significantly lower at certain clinical sites. Ultimately we believe that these differences did not significantly affect our results since this study focused on genotype-phenotype associations. Finally, despite our best efforts, we did not replicate the other GWAS signals. This may be because many of the replication cohorts were of primarily European ancestry, the original trials were only powered to detect a difference in their primary endpoint, and the replication cohorts had a different outcome definition (change in HbA1c at 1 year rather than the time-to-event analyses conducted in GRADE).

In summary, we report the results of a large pharmacogenomic study conducted in a randomized clinical trial, comparing four commonly prescribed medications to achieve or maintain glycemic control in addition to metformin. This study illustrates how the use of genetic data from clinical trials can enhance our understanding of pharmacogenomic response to glucose-lowering medications and contribute to the advancement of precision medicine.

Germline genetic predictors are unique biomarkers that are accessible, accurate, relatively cheap, and fixed for life. For pharmacogenetic precision medicine to take hold in public health, relevant genetic information needs to become available at the population level, as *de novo* genotyping for each indication would become financially unsustainable. Larger and better-powered studies are needed to generate data that identifies the most effective medication for each patient based on genotype.

## METHODS

### GRADE Study design

The GRADE Study is an U.S. randomized controlled trial in 5,047 participants from diverse ancestries with <10 years duration of diagnosed T2D and an HbA1c of 6.8-8.5% (50.8-69.4 mmol/mol) on metformin monotherapy. Participants were consented and enrolled at 36 clinical centers with 9 additional sub-sites in the U.S. and randomized to one of four glucose-lowering medications: insulin glargine, glimepiride, liraglutide, or sitagliptin.^27^

### Genome-wide genotyping and genotype imputation

A total of 4,749 participants, of whom >30% self-reported as non-White, consented to genetic testing on the Illumina Global Screening Array v3.0 chip, which contains >730,000 markers that provide genome-wide and exonic coverage. After excluding samples with poor DNA quality, 4,701 samples underwent genome-wide genotyping, and genotype data were successfully obtained for 4,689 participants. In the first stage of quality control (QC), GenomeStudio was utilized to perform raw data processing, including automatic clustering and QC based on GenTrain score, cluster separation, and Mendelian and replication errors.

Subsequently, using PLINK,^28^ we performed variant-based QC, excluding variants with i) missing call rate ≥0.05, ii) significant deviation from Hardy-Weinberg equilibrium (HWE) *p* ≤1×10^−5^ and iii) minor allele frequency (MAF) <0.01. Then we employed the following filters for sample-based QC: i) gender discordance between the self-reported and genetically predicted sex, ii) missing data rates or outlying heterozygosity rate, iii) subject relatedness, and iv) identification of individuals of divergent ancestry. After applying all quality control criteria, the final dataset included 4,572 participants with high-quality genotype data. A two-stage imputation procedure was performed. First, pre-phasing of the genotypes to whole-chromosome haplotypes was performed with SHAPEIT2.^29^ Imputation was performed with the TOPMed Imputation Server using the TOPMed reference panel.^30^ Genome annotations were generated using the GRCh38 assembly. The study was approved by the MGB IRB (#2023P000842).

### Pharmacogenetic GWAS in GRADE

We conducted separate GWAS survival analyses within each treatment group (glargine, glimepiride, liraglutide, and sitagliptin) using the gwasurvivr package.^31^ The primary and secondary metabolic outcomes were defined as an HbA1c ≥7% (53.0 mmol/mol) and >7.5% (58.5 mmol/mol) respectively, both subsequently confirmed. Cox proportional hazards (PH) regression models for time to event were used to characterize the nature of the association of candidate single nucleotide polymorphisms (SNPs) with the primary and secondary metabolic outcomes as a function of allele count, adjusting for baseline HbA1c, body mass index (BMI), age at randomization, sex, and 10 ancestry principal components. All ancestry groups were pooled for each GWAS within each treatment group. For the GWAS, genetic variants were filtered to a minor allele count ≥5 for individuals who reached the event and for those who did not. For reporting, we only considered variants with available results for both the primary and secondary outcome, though initial Manhattan plots (Figure S1) contain GWAS significant findings for each outcome separately. Variants were filtered to imputation quality R² ≥0.80. Analyses was performed under the intention-to-treat principle, with sensitivity analyses in an as-treated subgroup.

Kaplan-Meier curves represented the cumulative incidence of the primary outcome by allele for genome-wide significant loci within each treatment group. Estimates of the additive effects of variant alleles using PH models were stratified by sex and adjusted as above to calculate hazard ratios and robust confidence limits. We also reported differences in the restricted mean survival times (RMST) over 4 years of follow-up. For top genetic variants that were located in geographic proximity, we performed a conditional analysis to assess for independence of these variants. In practice, when two variants in a genomic region were strongly associated with the glycemic outcome, we tested a separate regression model that included both variants and assessed whether the association of each of the variants was still significant when both included in the regression model. We used a similar approach to express differences between treatment groups within allele subgroup. Analyses were conducted in R versions 4.1.1 and 4.2.1.

### Replication of GRADE GWAS findings

We sought replication of top findings that emerged from our sulfonylurea GWAS in GRADE by using the published GWAS meta-analysis by Dawed *et al.*,^5^ which comprised 5,485 White Europeans with T2D. The primary outcome of the study was HbA1c reduction (baseline minus follow-up) after 12 months of therapy with a sulfonylurea. To validate GWAS findings from the liraglutide arm in GRADE, we performed a look-up of top genetic variants in the published pharmacogenomic study of GLP-1 receptor agonists,^7^ which included 4,571 individuals with T2D from diverse ancestries. Glycemic response was defined as a quantitative trait of HbA1c reduction after 6 months of therapy with a GLP-1 receptor agonist.

### Replication in ORIGIN trial

For top findings in the glargine GWAS in GRADE, we explored their association with glycemic outcomes in the ORIGIN trial, which examined the effect of 1) titrated basal insulin glargine versus standard care and 2) n-3 fatty-acid supplements versus placebo on cardiovascular outcomes.^8^ In ORIGIN, a total of 12,537 individuals with cardiovascular risk factors and dysglycemia (impaired fasting glucose, impaired glucose tolerance, or type 2 diabetes) were randomized to glargine insulin versus standard care and n-3 fatty acids versus placebo using a 2 × 2 factorial design. They were followed for a median of 6.2 years for development of cardiovascular outcomes. In ORIGIN, genotyping was performed in 4,390 individuals using the HumanCoreExome Beadchip-12 v1.0 and v1.1 (Illumina), which measured 551,839 markers.^32^

Imputation was performed using the TOPMed-r2 reference panel. Of the ORIGIN participants with genetic data, we restricted the analyses to individuals with T2D, which totaled 4,015 participants (1,709 Europeans, 2,073 Latin Americans, 233 Africans). A total of 1,993 of these participants were randomized to the glargine arm (+/- n-3 fatty acids).

In participants with T2D randomized to the glargine treatment arm of ORIGIN, we performed a GWAS of the 1-year change in HbA1c. A multiple linear regression model tested allelic associations with 1-year change in HbA1c, assuming an additive model. One-year change was defined as one-year minus baseline value. Analyses were adjusted for age, sex, baseline BMI, first 10 ancestry principal components, and the baseline HbA1c. Because participants in ORIGIN may have additionally been on metformin, sulfonylurea, or thiazolidinedione treatment, a covariate was added to assess for presence or absence of a diabetes co-medication at baseline. Analyses were conducted separately by ancestry (European [n=819] and Latin Americans [n=931]) and then meta-analyzed thereafter. PLINK version 1.9 was used.^33^ GWAS results were filtered to a study-wide MAF>1% and imputation quality >0.3. We set a significance threshold of *p*<0.008 (0.05/6 independent variants).

### Replication in Outcome Reduction in SAVOR-TIMI 53 trial

SAVOR-TIMI 53 was a randomized, double-blind, placebo-controlled trial to assess the cardiovascular efficacy and safety of saxagliptin, a DPP4-inhibitor, in 16,492 participants with T2D and history of, or at elevated risk of, cardiovascular events.^9^ They were followed for a median of 2.1 years, and the primary endpoint was a composite outcome of cardiovascular death, myocardial infarction, or ischemic stroke. In this cohort, HbA1c was systematically measured at baseline, at months 12, 24, 36, and at the final study visit. A total of 8,548 participants who consented for genetic analysis underwent genome-wide genotyping using the Illumina Multi-Ethnic Genotyping Array. Imputation was performed with the TOPMed Freeze5 reference panel. Details of genotyping and imputation have been described in previous publications.^34^ Of these participants, 50.3% (n=4,299) received saxagliptin and 49.7% (n=4,249) received placebo. 12.2% of individuals did not have HbA1c measurements at 12 months and were excluded (n=523). For analyses, we restricted our analyses to individuals with genetic data and available HbA1c measurements in the saxagliptin arm (n=3,776).

In individuals randomized to saxagliptin, we performed a GWAS of the 1-year change in HbA1c. A multiple linear regression model tested associations between genetic variation and the outcome of 1-year change in HbA1c (calculated as follow-up HbA1c at 12 months minus baseline HbA1c), assuming an additive model. Ancestry groups were analyzed together and 10 global PCs were constructed for use in model adjustment. Additional covariates included age, sex, BMI, baseline HbA1c, and additional variables that represented concomitant use of additional glucose-lowering medications (metformin, sulfonylurea, thiazolidinedione, or insulin). REGENIE version 3.4.1 was used for analysis. GWAS results were filtered to MAF >0.1% and imputation quality R^2^ >0.3. We used a significance threshold of *p*<0.017 (0.05/3 independent variants).

### Assessment of **β**-cell function utilizing serial OGTTs in GRADE

To gain physiologic insight into the influence of the variant on β-cell response, we plotted the results of serial oral glucose tolerance tests (OGTTs) at years 0, 1, and 3 post-randomization and examined the association of top variants with fasting glucose, fasting C-peptide, and C-peptide index (CPI) using a separate robust M-estimate regression model at each year with a Huber Ψ function and adjusting by 10 ancestry principal components. The CPI was calculated as the increment in C-peptide above fasting (0 min) over the first 30 min of the OGTT divided by the increment in glucose over the same period: 100(C_30_ − C_0_)/(G_30_ − G_0_).^35^ In addition to an intention-to-treat analysis, we also performed an as-treated analysis, which excluded 1558 OGTTs obtained from participants who had either never taken their randomly assigned medication, or had deviated from their assigned regimen before the OGTT.

### Assessment of glimepiride response variants in SUGAR-MGH

The Study to Understand the Genetics of the Acute Response to Metformin and Glipizide in Humans (SUGAR-MGH) is a pharmacogenetic study in which 1,000 individuals naïve to type 2 diabetes (T2D) medications provided DNA for genome-wide genotyping and received a single 5 mg dose of glipizide, followed by a short course of metformin and an OGTT.^10,11^ We applied a mixed-effects model with a random effect for individuals, using the lme4 package and including a time × variant interaction term, to evaluate the influence of the top glimepiride-response GWAS variant on glucose levels and log-transformed insulin measurements over 240 minutes following glipizide administration. Models were adjusted for age, sex, BMI, and 10 ancestry principal components.

### Effect of CRISPR/Cas9-induced mutations in *slc2a2* on glucose content in glimepiride-treated zebrafish larvae

#### Zebrafish husbandry

Adult AB zebrafish were maintained in tanks in a system with re-circulating filtered water (Tecniplast, Buguggiate, Italy), at five fish/L and 28°C. Fish were fed robotically twice daily with dry food (Sparos, Olhao, Portugal) and once daily with rotifers. Adults and larvae were raised and maintained under a 14h - 10h light - darkness cycle with a 30 min dimming / lighting period in between. Adult fish were crossed to generate offspring for experiments.

#### Target design and gRNA/Cas9 complex preparation

To target the zebrafish orthologue of *SLC2A2 (slc2a2*, ENSDARG00000056196), we used the same single guide RNA (gRNA) that recently showed high on target efficiency in a systematic genetic screen for effects on T2D traits (submitted for publication). The gRNA (GTCGCCATCTTCTCTGTCGG) was designed using CRISPOR v4.99 and selected based on its estimated efficiency and specificity, i.e., a high MIT specificity score, high cutting Frequency Determination score, no predicted off-target activity sites, and no overlap with known SNPs.^36^ PCR primers flanking the gRNA region were designed using CRISPOR (forward: tgtaaaacgacggccagtGTCATTGAGCGGCACTACG; reverse: gtgtcttTTGCCTTTGAGCCTTTCCAG) and acquired from Integrated DNA Technologies (Coralville, Iowa, USA).

#### Micro-injections

To obtain a 50 μM duplex for micro-injection, we mixed equal volumes of 100 μM tracrRNA and 100 μM single gRNA. We next denatured the oligo at 95 °C for 5 min, followed by incubation at a gradually decreasing temperature (0.1 °C/sec), from 95 °C to 25 °C, and finally 5 min at 25 °C. We then combined the duplex with Alt-R® S.p. Cas9 nuclease, v. 3 (IDT) and added water to achieve a final concentration of 5 μM. As a visual guide for microinjections, we added 1 μM of 0.5% phenol red (Santa Ana, USA). We micro-injected 300 pmol of the CRISPR/Cas9 complex per egg, at the single cell stage, in >1000 fertilized zebrafish eggs per crossing. We used a stereomicroscope with eggs placed in an agar plate with microchannels.

Eggs targeted at *slc2a2* (from here on referred to as ‘crispants’) as well as sibling controls were micro-injected with a gRNA targeting exon 4 of *kita*, to ensure all larvae included in the experiment underwent micro-injections at the single cell stage, DNA editing, and DNA repair. Crispants and controls were micro-injected with the same absolute amount of gRNA and Cas9. *Kita* is essential for melanocyte migration, so successfully edited larvae have less pigmentation throughout embryonic development. Micro-injections were repeated three times (3 batches) to accomplish the final sample size.

#### Raising larvae from day 0 to 10

We raised micro-injected eggs in an incubator at 28.5°C with 0.01% methylene blue in filtered water until day 5. On the afternoon of day 5, we optically identified larvae that were successfully micro-injected and edited at day 0 using absence of pigment^23^. These larvae were transferred to tanks at a density of 30 larvae in 300 ml of filtered water per 1 L tank, at a fixed ratio of 50% crispants and 50% controls. Based on the number of crispants and controls available on day 5, we raised 8, 9 and 7 tanks of larvae per batch. Larvae were fed 16.29 mg of dry food per tank twice per day, from the afternoon of day 5 until the afternoon of day 9. On days 8 and 9, we added glimepiride (Cayman Chemicals, Ann Arbor, Michigan, USA; solubility: 3 mg/ml) dissolved in DMSO to a final concentration of 50 μM glimepiride and 0.82% DMSO in 300 ml of water to 4, 5, and 4 tanks in batches 1-3. The other tanks were only supplemented with the same amount of DMSO. Water was exchanged in the morning of days 8 – immediately before first adding the drug – and 9 to avoid bacterial growth. Glimepiride was replenished after the water change on day 9. The dose of glimepiride used was based on the results of a pilot study, in which 50 μM resulted in 0.59 ± 0.22 SD units lower glucose content (P = 0.008) than observed in untreated sibling controls (n = 47, Figure S7). On the morning of day 10, larvae were sacrificed by exposure to tricaine (3.06 µM) and placed on ice for 5 min. Next, larvae were washed twice with PBS, placed in 96-well plates at one larva per well, and frozen at −20 °C for later processing.

#### Sample prep and random glucose and protein content

We homogenized larvae individually by adding two 1.4 mm zirconium beads (Diagnostics, NJ, USA) into each well, together with 240 µl of ice-cold PBS. Each plate was homogenized using a SPEX SamplePrep 1600 MiniG (Gammadata Instruments, Uppsala, Sweden) for 2 min at 1000 rpm. The homogenized larvae were centrifuged at 3500 g for 5 min at 4 °C. Glucose content was quantified in 10 µl of supernatant using Glucose-Glo™ (J6022, Promega, WI, USA) and following the manufacturer’s instructions. Protein content was assessed using the Pierce bicinchoninic acid (BCA) Protein Assay Kit (Thermo Fisher Scientific, Waltham, MA, USA). For glucose content, luminescence was measured on a VARIOSKAN LUX plate (ThermoFisher, Waltham, MA, USA) and technical replicates were performed and averaged. Protein content was measured by absorbance at 562 nm using the same device.

#### Genotyping for CRISPR/Cas9-induced mutations in slc2a2

We extracted DNA from the residual of homogenized tissue using Proteinase K (Sigma-Aldrich, Munich, Germany) and lysis buffer (10 mM Tris-HCl pH8, 50 mM KCl, 0.2 mM EDTA, 0.3% Tween 20, 0.3% Igepal). M13 tailed forward and pigtailed reverse primers were selected flanking the CRISPR/Cas9-targeted region. A PCR reaction was set up using a final concentration of 0.25 units per 10 µl of OneTaq® DNA Polymerase (M0480) (New England Biolabs, Ipswich, MA, USA), 1X OneTaq Standard Reaction Buffer, 0.1 μM forward primer, 0.2 μM reverse primer, 0.2 μM fluorescently labeled M13, 0.2 μM dNTPs and >2 ng of unpurified extracted DNA and water to reach a final reaction volume of 10 µl. PCR steps were as follows: 30 sec at 94 °C; 34 cycles of 30 sec at 94 °C, 45 sec of annealing at 58 °C, 30 sec at 68 °C; a final extension step at 69 °C; followed by a hold at 4 °C.

PCR products were diluted 5 times and 1.5 μl was transferred to a barcoded 96-well plate pre-loaded with 10 μl of Hi-Di™ formamide (ThermoFisher, Waltham, USA) and GeneScan™ - 400HD ROX™. DNA from 16 un-injected sibling controls was added to each plate. Size standards and Hi-Di only wells were loaded on each plate to allow DNA fragment size identification and visualization of PCR product vs. background noise. Assembled barcoded 96-well plates were heated for 5 min at 95 °C and cooled on ice. Finally, plates were loaded into an Applied Biosystems® 3730XL DNA analyzer (Applied Biosystems, Waltham, USA) for capillary electrophoresis.

We analyzed results using Peak Scanner version 2.0 (ThermoFisher Scientific) and a custom-written script in R (v.4.1.0) to perform quality control and determine the (proportional) area of wild type (WT) and non-WT peaks per sample. Larvae were considered mutated at *slc2a2* if ≤30% of the peak area was of wild-type length; and controls if >85% of the peak area was of wild-type length.

#### Data analysis

Within each batch, survival from day 1 – after removing dead and unfertilized eggs – to day 5 was similar in crispants targeted at *slc2a2* and *kita* (56%, 74%, 66% in batches 1-3) vs. sibling controls only targeted at *kita* (64%, 77%, 68%) (*p*_t-test_ = 0.52). Across all three batches combined, day 1-to-5 survival rates of 61% in *slc2a2* crispants and 68% in controls are similar to those we observed earlier across six other T2D candidate genes micro-injected with the same amount of gRNA and Cas9 (57 ± 13%). On average, 16.3 ± 4.6 larvae per tank survived from day 5 to day 10, resulting in a total sample of 390 larvae at day 10 (132, 169 and 89 larvae per batch, or 54%). Survival was similar in glimepiride treated (16.9 ± 5.1 larvae per tank, 56%) and untreated tanks (15.5 ± 4.1, 52%, *p*_t-test_ = 0.45). Survival from day 5 to 10 was also similar in treated and untreated larvae compared with crispants for the previously mentioned other T2D candidate genes (50 ± 19%).

Within each batch, glucose content was first set to missing if duplicates differed by >25%. Next, glucose and protein values outside the mean ± 2.5 × SD range were set to missing. These steps excluded 39 larvae from the analysis (1.6 ± 1.7 larvae per tank, *p*_t-test_ treated vs. untreated = 0.66). Finally, glucose content was inverse-normally transformed within each batch to a mean of 0 and an SD of 1.

To examine if the effect of glimepiride treatment on glucose content was the same with/without CRISPR/Cas9-induced mutations in *slc2a2*, we used a multiple linear regression analysis with an interaction term, adjusting for batch and protein content. Adjusting for batch and/or protein content did not affect conclusions about the main treatment effect or the interaction between treatment and pLOF mutations in *slc2a2*. Data management and statistical analysis were performed in Stata MP v16.1.

#### Ethical permits

All animal care taking routines and experimental procedures described here have been approved by the Uppsala Board for Animal Research (Djurförsöksetiska nämnd) of the Swedish ministry of agriculture (Jordbruksverket) under permit number Dnr 5.8.18-13680/2020.

## Data Availability

The summary statistics from this study will be available at the Common Metabolic Diseases Knowledge Portal (https://hugeamp.org) and the GWAS Catalog (https://www.ebi.ac.uk/gwas/) following article publication. The Glycemia Reduction Approaches in Diabetes: A Comparative Effectiveness Study (GRADE) is funded by the National Institute of Diabetes and Digestive and Kidney Diseases (NIDDK). This manuscript is based on follow-up data and outcome assessments from the 5047 participants enrolled into the study. This database is available in the NIDDK Central Repository (https://repository.niddk.nih.gov/study/151).

## ACKNOWLEDGEMENTS

**Funding.** This work was supported by NIDDK grant R01 DK123019 to J.C.F. Genotyping was supported by a grant from the Polish Ministry of Health. J.H.L. is supported by NIDDK K23 DK131345. L.S. is supported by funds from the American Diabetes Association grant 11-22-PDFPM-03 and the National Institute of Diabetes and Digestive and Kidney Diseases of the National Institutes of Health under Award Number U01 DK140757. L.S., A.C-R., P.K., M.C., A.K. are supported by funds from the Ministry of Education and Science of Poland within the project “Excellence Initiative - Research University,” the Ministry of Health of Poland within the project “Center of Artificial Intelligence in Medicine at the Medical University of Bialystok,” and the Medical Research Agency within the project “Regional Center for Digital Medicine at the Medical University of Bialystok” (grant number 2023/ABM/02/00008). A.H.-C. is supported by the American Diabetes Association grant 11-23-PDF-35. A.L. is supported by grant 2020096 from the Doris Duke Foundation and the American Diabetes Association Grant 7-22-ICTSPM-23. J.M.M. is supported by American Diabetes Association grant #11-22-ICTSPM-16 and by NHGRI U01HG011723, by the National Institute of Diabetes and Digestive and Kidney Diseases of the National Institutes of Health under Award Number R01DK137993 and U01 DK140757, AMP CMD award from RFP 6 from the Foundation for the National Institutes of Health, and a Medical University of Bialystok (MUB) grant from the Ministry of Science and Higher Education (Poland).

The GRADE Study was supported by a grant (U01DK098246) from the National Institute of Diabetes and Digestive and Kidney Diseases (NIDDK) of the National Institutes of Health (NIH); a U34 planning grant (U34-DK-088043) for the planning of the trial from the NIDDK; funding for the initial planning meeting regarding the U34 proposal from the American Diabetes Association; the National Heart, Lung, and Blood Institute; the Centers for Disease Control and Prevention; resources and facilities from the Department of Veterans Affairs; grants (P30 DK017047, P30 DK020541–44, P30 DK020572, P30 DK072476, P30 DK079626, P30 DK092926, U54 GM104940, UL1 TR000170, UL1 TR000439, UL1 TR000445, UL1 TR001102, UL1 TR001108, UL1 TR001409, 2UL1TR001425, UL1 TR001449, UL1 TR002243, UL1 TR002345, UL1 TR002378, UL1 TR002489, UL1 TR002529, UL1 TR002535, UL1 TR002537, UL1 TR002541, and UL1 TR002548) from the NIH; educational materials from the National Diabetes Education Program; and donated medications and supplies from Becton Dickinson, Bristol Myers Squibb, Merck, Novo Nordisk, Roche Diagnostics, and Sanofi. The zebrafish study / M.d.H. is supported by the Swedish Heart-Lung Foundation (20230518), the Swedish Research Council (2023-02556), and the NIH/NIDDK-funded Accelerating Medicines Partnership for Common Metabolic Disorders (5UM1DK105554-5000826-5500002718).

## Competing interests

J.H.L. has received speaker honoraria from Jubilant Therapeutics, Inc. H.L. is currently employed at Astra Zeneca as an RWE Associate Director. A.L. had a financial interest, during a portion of this grant, in Merck & Co., Inc., a company whose medication is being evaluated in this research. A.L.’s interests were reviewed and are managed by Massachusetts General Hospital and Mass General Brigham in accordance with their conflict of interest policies. H.C.G. holds the McMaster-Sanofi Population Health Institute Chair in Diabetes Research and Care. He reports research grants from Eli Lilly, Novo Nordisk, and Hanmi Pharmaceutical; grants to support continuing education programs from Eli Lilly, Abbott, Sanofi, Novo Nordisk, and Boehringer Ingelheim; honoraria for speaking from AstraZeneca, Eli Lilly, Zuellig, and Jiangsu Hanson; and consulting fees from Abbott, Bayer, Biolinq, Eli Lilly, Novo Nordisk, Pfizer, Shionogi, and Zealand. D.L.B. discloses the following relationships - Advisory Board: Angiowave, Antlia Bioscience, Bayer, Boehringer Ingelheim, CellProthera, Cereno Scientific, E-Star Biotech, High Enroll, Janssen, Level Ex, McKinsey, Medscape Cardiology, Merck, NirvaMed, Novo Nordisk, Repair Biotechnologies, Stasys, Tourmaline Bio; Board of Directors: American Heart Association New York City, Angiowave (stock options), Bristol Myers Squibb (stock), DRS.LINQ (stock options), High Enroll (stock); Consultant: Alnylam, Altimmune, Broadview Ventures, Corcept Therapeutics, Corsera, GlaxoSmithKline, Hims, SERB, SFJ, Summa Therapeutics, Worldwide Clinical Trials; Data Monitoring Committees: Acesion Pharma, Assistance Publique-Hôpitaux de Paris, Baim Institute for Clinical Research, Boston Scientific (Chair, PEITHO trial), Cleveland Clinic, Contego Medical (Chair, PERFORMANCE 2), Duke Clinical Research Institute, Mayo Clinic, Mount Sinai School of Medicine (for the ABILITY-DM trial, funded by Concept Medical; for ALLAY-HF, funded by Alleviant Medical), Novartis, Population Health Research Institute; Rutgers University (for the NIH-funded MINT Trial); Honoraria: American College of Cardiology (Senior Associate Editor, Clinical Trials and News, ACC.org; Chair, ACC Accreditation Oversight Committee), Arnold and Porter law firm (work related to Sanofi/Bristol-Myers Squibb clopidogrel litigation), Baim Institute for Clinical Research (AEGIS-II executive committee funded by CSL Behring), Belvoir Publications (Editor in Chief, Harvard Heart Letter), Canadian Medical and Surgical Knowledge Translation Research Group (clinical trial steering committees), CSL Behring (AHA lecture), Duke Clinical Research Institute, Engage Health Media, HMP Global (Editor in Chief, Journal of Invasive Cardiology), Medtelligence/ReachMD (CME steering committees), MJH Life Sciences, Oakstone CME (Course Director, Comprehensive Review of Interventional Cardiology), Philips (Becker’s Webinar on AI), Population Health Research Institute, WebMD (CME steering committees), Wiley (steering committee); Other: Clinical Cardiology (Deputy Editor); Progress in Cardiovascular Diseases (Deputy Editor); Patent: Sotagliflozin (named on a patent for sotagliflozin assigned to Brigham and Women’s Hospital who assigned to Lexicon; neither I nor Brigham and Women’s Hospital receive any income from this patent); Research Funding: Abbott, Acesion Pharma, Afimmune, Alnylam, Amarin, Amgen, AstraZeneca, Atricure, Bayer, Boehringer Ingelheim, Boston Scientific, CellProthera, Cereno Scientific, Chiesi, Cleerly, CSL Behring, Faraday Pharmaceuticals, Fractyl, Idorsia, Janssen, Javelin, Lexicon, Lilly, Medtronic, Merck, MiRUS, Moderna, Novartis, Novo Nordisk, Pfizer, PhaseBio, Regeneron, Reid Hoffman Foundation, Roche, Sanofi, Stasys, 89Bio; Royalties: Elsevier (Editor, Braunwald’s Heart Disease); Site Co-Investigator: Cleerly. N.A.M. reports funds through a National Institutes of Health grant (K08HL153950) and is involved in clinical trials with Amgen, Pfizer, Ionis, Novartis, and AstraZeneca without personal fees, payments, or increase in salary.

C.T.R. reports research grant through institution: Anthos, AstraZeneca, Daiichi Sankyo, Janssen and Novartis; Honoraria for scientific advisory boards and consulting: Anthos, Bayer, Bristol Myers Squibb, Daiichi Sankyo, Janssen, Pfizer. M.S.S. reports research grant support through Brigham and Women’s Hospital from: Abbott; Amgen; Anthos Therapeutics, Inc.; AstraZeneca; Boehringer Ingelheim; Daiichi-Sankyo; Ionis; Marea; Merck; Novartis; Pfizer; Saghmos Therapeutics; Verve Therapeutics. Consulting for: Amgen; AMPEL BioSolutions; Anthos Therapeutics, Inc.; AstraZeneca; Beren Therapeutics; Boehringer Ingelheim; CCRN; Dr. Reddy’s Laboratories; General Medicines; Merck; NATF; Novo Nordisk; Precision BioSciences. Y.P.L., N.A.M., C.T.R., and M.S.S. are members of the TIMI (Thrombolysis in Myocardial Infarction) Study Group, which has received institutional research grant support through Brigham and Women’s Hospital from Abbott, Abiomed, Inc., Amgen, Anthos Therapeutics, ARCA Biopharma, Inc., AstraZeneca, Boehringer Ingelheim, Daiichi-Sankyo, Ionis Pharmaceuticals, Inc., Janssen Research and Development, LLC, MedImmune, Merck, Novartis, Pfizer, Regeneron Pharmaceuticals, Inc., Roche, Saghmos Therapeutics, Inc., Softcell Medical Limited, The Medicines Company, Verve Therapeutics, Inc., Zora Biosciences. E.R.P. has received honoraria for speaking from Novo Nordisk and Eli Lilly. J.C.F. has received consulting honoraria from Alveus Therapeutics. J.C.F. and M.d.H. are PIs on different research projects funded by Novo Nordisk. L.S., M.T., S.N., E.M., A.E., T.Y.W. A.C.-R., P.K., M.C., A.H.-C., M.V. J.B.M., M.Y.N. R.J.F.L., M.P., F.A.M., A.D., L.S.S., A.J.K., S.K., N.Y., and J.M.M. have nothing to disclose.

## Author Contributions

J.H.L., L.S., N.Y., J.M.M., and J.C.F. conceived and designed the study. Genotyping was performed by L.S., A.C.-R. P.K., M.C., and A.K. Quality control, imputation of the genetic data, and GWAS analyses and interpretation in GRADE were performed by J.H.L., L.S., M.T., H.L., A.L., S.E.K., N.Y., J.M.M., and J.C.F. J.H.L. and S.N. performed statistical analysis in SUGAR-MGH. M.P., H.C.G., F.A.M., Y.L., D.L.B., N.A.M., C.T.R., M.S.S., A.Y.D., and E.R.P. performed the replication analysis in their respective cohorts. E.M., A.E., M.d.H. conducted the zebrafish experiments. J.H.L., L.S., and J.M.M. drafted the initial manuscript. All authors participated in interpretation of the results, provided critical review of the manuscript, and approved the manuscript to be submitted for publication. J.M.M. and J.C.F. are the guarantors of this work.

## Prior Presentation

Portions of this study were previously presented at the 83^rd^ Scientific Sessions of the American Diabetes Association in San Diego, CA, June 23-26, 2023, and the 59^th^ Annual Meeting of the European Association for the Study of Diabetes in Hamburg, Germany, October 2-6, 2023.

## Code availability

The study utilized previously published analysis tools as described in the Methods.

## Notes

### Clinical Trial

NCT01794143

### Author Declarations

The study was approved by the MGB IRB (#2023P000842).

## REFERENCES

1. Kahn SE, Haffner SM, Heise MA, et al. Glycemic durability of rosiglitazone, metformin, or glyburide monotherapy. N Engl J Med 2006;355(23):2427–43. DOI: 10.1056/NEJMoa066224.

2. Nathan DM, Buse JB, Kahn SE, et al. Rationale and design of the glycemia reduction approaches in diabetes: a comparative effectiveness study (GRADE). Diabetes Care 2013;36(8):2254–61. DOI: 10.2337/dc13-0356.

3. Grade Study Research Group, Nathan DM, Lachin JM, et al. Glycemia Reduction in Type 2 Diabetes - Glycemic Outcomes. N Engl J Med 2022;387(12):1063–1074. DOI: 10.1056/NEJMoa2200433.

4. Zhou K, Donnelly L, Yang J, et al. Heritability of variation in glycaemic response to metformin: a genome-wide complex trait analysis. Lancet Diabetes Endocrinol 2014;2(6):481–7. DOI: 10.1016/S2213-8587(14)70050-6.

5. Dawed AY, Yee SW, Zhou K, et al. Genome-Wide Meta-analysis Identifies Genetic Variants Associated With Glycemic Response to Sulfonylureas. Diabetes Care 2021;44(12):2673–2682. DOI: 10.2337/dc21-1152.

6. Zhou K, Yee SW, Seiser EL, et al. Variation in the glucose transporter gene SLC2A2 is associated with glycemic response to metformin. Nat Genet 2016;48(9):1055–1059. DOI: 10.1038/ng.3632.

7. Dawed AY, Mari A, Brown A, et al. Pharmacogenomics of GLP-1 receptor agonists: a genome-wide analysis of observational data and large randomised controlled trials. Lancet Diabetes Endocrinol 2023;11(1):33–41. DOI: 10.1016/S2213-8587(22)00340-0.

8. Origin Trial Investigators, Gerstein HC, Bosch J, et al. Basal insulin and cardiovascular and other outcomes in dysglycemia. N Engl J Med 2012;367(4):319–28. DOI: 10.1056/NEJMoa1203858.

9. Scirica BM, Bhatt DL, Braunwald E, et al. Saxagliptin and cardiovascular outcomes in patients with type 2 diabetes mellitus. N Engl J Med 2013;369(14):1317–26. DOI: 10.1056/NEJMoa1307684.

10. Walford GA, Colomo N, Todd JN, et al. The study to understand the genetics of the acute response to metformin and glipizide in humans (SUGAR-MGH): design of a pharmacogenetic resource for type 2 diabetes. PLoS One 2015;10(3):e0121553. DOI: 10.1371/journal.pone.0121553.

11. Li JH, Brenner LN, Kaur V, et al. Genome-wide association analysis identifies ancestry-specific genetic variation associated with acute response to metformin and glipizide in SUGAR-MGH. Diabetologia 2023;66(7):1260–1272. DOI: 10.1007/s00125-023-05922-7.

12. Broadaway KA, Brotman SM, Rosen JD, et al. Liver eQTL meta-analysis illuminates potential molecular mechanisms of cardiometabolic traits. Am J Hum Genet 2024;111(9):1899–1913. DOI: 10.1016/j.ajhg.2024.07.017.

13. van de Bunt M, Manning Fox JE, Dai X, et al. Transcript Expression Data from Human Islets Links Regulatory Signals from Genome-Wide Association Studies for Type 2 Diabetes and Glycemic Traits to Their Downstream Effectors. PLoS Genet 2015;11(12):e1005694. (In eng). DOI: 10.1371/journal.pgen.1005694.

14. Alonso L, Piron A, Moran I, et al. TIGER: The gene expression regulatory variation landscape of human pancreatic islets. Cell Rep 2021;37(2):109807. DOI: 10.1016/j.celrep.2021.109807.

15. Sun B, Chen H, Xue J, Li P, Fu X. The role of GLUT2 in glucose metabolism in multiple organs and tissues. Mol Biol Rep 2023;50(8):6963–6974. DOI: 10.1007/s11033-023-08535-w.

16. Suzuki K, Hatzikotoulas K, Southam L, et al. Genetic drivers of heterogeneity in type 2 diabetes pathophysiology. Nature 2024;627(8003):347–357. DOI: 10.1038/s41586-024-07019-6.

17. Lagou V, Jiang L, Ulrich A, et al. GWAS of random glucose in 476,326 individuals provide insights into diabetes pathophysiology, complications and treatment stratification. Nat Genet 2023;55(9):1448–1461. DOI: 10.1038/s41588-023-01462-3.

18. Sinnott-Armstrong N, Tanigawa Y, Amar D, et al. Genetics of 35 blood and urine biomarkers in the UK Biobank. Nat Genet 2021;53(2):185–194. DOI: 10.1038/s41588-020-00757-z.

19. Pearson ER, Flechtner I, Njolstad PR, et al. Switching from insulin to oral sulfonylureas in patients with diabetes due to Kir6.2 mutations. N Engl J Med 2006;355(5):467–77. DOI: 10.1056/NEJMoa061759.

20. Henquin JC. Triggering and amplifying pathways of regulation of insulin secretion by glucose. Diabetes 2000;49(11):1751–60. (In eng). DOI: 10.2337/diabetes.49.11.1751.

21. Henquin JC. The dual control of insulin secretion by glucose involves triggering and amplifying pathways in beta-cells. Diabetes Res Clin Pract 2011;93 Suppl 1:S27–31. DOI: 10.1016/S0168-8227(11)70010-9.

22. Al-Rubeaan K, Alsayed M, Ben-Nakhi A, et al. Characteristics and Treatment Patterns of Patients with Type 2 Diabetes Mellitus in the Middle East and Africa Cohort of the DISCOVER Study Program: a Prospective Study. Diabetes Ther 2022;13(7):1339–1352. DOI: 10.1007/s13300-022-01272-6.

23. Barber MJ, Gotham D, Bygrave H, Cepuch C. Estimated Sustainable Cost-Based Prices for Diabetes Medicines. JAMA Netw Open 2024;7(3):e243474. DOI: 10.1001/jamanetworkopen.2024.3474.

24. International Diabetes Federation. IDF Diabetes Atlas, 10th edn. (https://www.diabetesatlas.org).

25. Guidelines on second-and third-line medicines and type of insulin for the control of blood glucose levels in non-pregnant adults with diabetes mellitus. Geneva2018.

26. Polonsky WH, Skinner TC. Perceived Treatment Efficacy: An Overlooked Opportunity in Diabetes Care. Clinical Diabetes 2010;28(2).

27. Wexler DJ, Krause-Steinrauf H, Crandall JP, et al. Baseline Characteristics of Randomized Participants in the Glycemia Reduction Approaches in Diabetes: A Comparative Effectiveness Study (GRADE). Diabetes Care 2019;42(11):2098–2107. DOI: 10.2337/dc19-0901.

28. Purcell S, Neale B, Todd-Brown K, et al. PLINK: a tool set for whole-genome association and population-based linkage analyses. Am J Hum Genet 2007;81(3):559–75. DOI: 10.1086/519795.

29. Delaneau O, Zagury JF, Marchini J. Improved whole-chromosome phasing for disease and population genetic studies. Nat Methods 2013;10(1):5–6. DOI: 10.1038/nmeth.2307.

30. Taliun D, Harris DN, Kessler MD, et al. Sequencing of 53,831 diverse genomes from the NHLBI TOPMed Program. Nature 2021;590(7845):290–299. DOI: 10.1038/s41586-021-03205-y.

31. Rizvi A KE, Morgan M, Sucheston-Campbell L. gwasurvivr: an R package for genome wide survival analysis. (https://github.com/suchestoncampbelllab/gwasurvivr).

32. Shah HS, Gao H, Morieri ML, et al. Genetic Predictors of Cardiovascular Mortality During Intensive Glycemic Control in Type 2 Diabetes: Findings From the ACCORD Clinical Trial. Diabetes Care 2016;39(11):1915–1924. DOI: 10.2337/dc16-0285.

33. Chang CC, Chow CC, Tellier LC, Vattikuti S, Purcell SM, Lee JJ. Second-generation PLINK: rising to the challenge of larger and richer datasets. Gigascience 2015;4:7. DOI: 10.1186/s13742-015-0047-8.

34. Marston NA, Patel PN, Kamanu FK, et al. Clinical Application of a Novel Genetic Risk Score for Ischemic Stroke in Patients With Cardiometabolic Disease. Circulation 2021;143(5):470–478. DOI: 10.1161/CIRCULATIONAHA.120.051927.

35. Seltzer HS, Allen EW, Herron AL, Jr., Brennan MT. Insulin secretion in response to glycemic stimulus: relation of delayed initial release to carbohydrate intolerance in mild diabetes mellitus. J Clin Invest 1967;46(3):323–35. (In eng). DOI: 10.1172/jci105534.

36. Concordet JP, Haeussler M. CRISPOR: intuitive guide selection for CRISPR/Cas9 genome editing experiments and screens. Nucleic Acids Res 2018;46(W1):W242–W245. DOI: 10.1093/nar/gky354.

