## Supplemental File for "Common genetic variants near *SLC2A2* and glycemic response to glimepiride in the GRADE comparative effectiveness clinical trial"

### LIST OF INVESTIGATORS

**GRADE Research Group (April 30, 2021)**

**Designations:** Principal Investigator (PI); Co-Principal Investigator (Co-PI); Co-Investigator (Co-I); Study Coordinator (SC); Recruitment/Retention Coordinator (RC); Research Staff (RS)

**Current Clinical Centers**

**Albert Einstein College of Medicine:** Crandall, JP (PI); McKee, MD (Co-PI, past); Behringer-Massera , S (Co-I, past); Brown-Friday, J (SC); Xhori, E (RC, past); Ballentine-Cargill, K (RS); Duran, S (RS); Estrella, H (RS); Gonzalez de la torre, S (RS, past); Lukin, J (RS, past)

**Atlanta VA Medical Center:** Phillips, LS (PI); Burgess, E (Co-I); Olson, D (Co-I); Rhee, M (Co-I); Wilson, P (Co-I); Raines, TS (SC); Boers, J (SC); Costello, J (SC); Maher-Albertelli, M (SC); Mungara, R (SC); Savoye, L (SC, past); White, CA (SC); Gullett, C (SC; past); Holloway, L (SC, past); Morehead, F (SC, past); Person, S (SC, past); Sibymon, M (SC, past); Tanukonda, S (SC, past); Adams, C (RC, past); Ross, A (RC, past)

**Baylor College of Medicine:** Balasubramanyam, A (PI); Gaba, R (Co-I); Gonzalez Hattery, E (SC); Ideozu, A (SC); Jimenez, J (SC); Montes, G (RC); Wright, C (RS, past)

**Baylor Scott & White Research Institute (Baylor Research Institute):** Hollander, P (PI); Roe, E (Co-I, past); Jackson, A (SC); Smiley, A (SC); Burt, P (SC, past); Estrada, L (RS); Chionh, K (RS, past)

**Case Western Reserve University/Cleveland VA/MetroHealth Medical Center:** Ismail-Beigi, F (PI); Falck-Ytter, C (Co-PI); Sayyed Kassem, L (Co-PI); Sood, A (Co-PI, past); Tiktin, M (Co-I, SC); Kulow, T (SC); Newman, C (SC); Stancil, KA (SC); Cramer, B (SC, past); Iacoboni, J (SC, past); Kononets, MV (SC, past); Sanders, C (SC, past); Tucker, L (SC, past); Werner, A (SC, past); Maxwell, A (RS); McPhee, G (RS); Patel, C (RS); Colosimo, L (RS, past); Krol, A (RS, past)

**Columbia University Medical Center:** Goland, R (PI); Pring, J (SC); Alfano, L (SC); Kringas, P (SC, past); Hausheer, C (RC, past); Tejada, J (RC, past); Gumpel, K (RS, past); Kirpitch, A (RS, past); Schneier, H (RS, past)

**Duke University Medical Center:** Green, JB (PI); AbouAssi, H (Co-I); Chatterjee, R (Co-I); Feinglos, MN (Co-I, past); English Jones, J (SC, RC); Khan, SA (SC, RC); Kimpel, JB (SC, past); Zimmer, RP (SC, past); Furst, M (RC, past); Satterwhite, BM (RS); Thacker, CR (RS); Evans Kreider, K (RS, past)

**Indiana University:** Mariash, CN (PI); Mather, KJ (PI, past); Ismail, HM (Co-I); Lteif, A (Co-I, past); Mullen, M (SC); Hamilton, T (SC, past); Patel, N (SC, past); Riera, G (RC); Jackson, M (RC, past); Pirics, V (RC, past); Aguillar, D (RS, past); Howard, D (RS, past); Hurt, S (RS, past)

**International Diabetes Center:** Bergenstal, R (PI); Carlson, A (Co-I); Martens, T (Co-I); Johnson, M (SC); Hill, R (SC); Hyatt, J (SC); Jensen, C (SC); Madden, M (SC); Martin, D (SC); Willis, H (SC); Konerza, W (RS); Yang, S (RS); Kleeberger, K (RS, past); Passi, R (RS, past)

**Kaiser Permanente Northwest:** Fortmann, S (PI); Herson, M (Co-I); Mularski, K (Co-I); Glauber, H (Co-I, past); Prihoda, J (Co-I, past); Ash, B (SC); Carlson, C (SC); Ramey, PA (SC); Schield, E (SC); Torgrimson-Ojerio, B (SC); Arnold, K (SC, past); Kauffman, B (SC, past); Panos, E (SC, past); Sahnow, S (RC); Bays, K (RS); Berame, K (RS); Cook, J (RS); Ghioni, D (RS); Gluth, J (RS); Schell, K (RS); Criscola, J (RS, past); Friason, C (RS, past); Jones, S (RS, past); Nazarov, S (RS, past)

**Kaiser Permanente of Georgia:** Barzilay, J (PI); Rassouli, N (Co-PI); Puttnam, R (Co-I); Ojoawo, B (SC); Nelson, R (RC); Curtis, M (SC, past); Hollis, B (SC, past); Sanders-Jones, C (SC, past); Stokes, K (SC, past); El-Haqq, Z (RS, past); Kolli, A (RS, past); Tran, T (RS, past)

**Massachusetts General Hospital:** Wexler, D (PI); Larkin, ME (Co-I, SC); Meigs, J (Co-I); Chambers, B (SC, past); Dushkin, A (SC, past); Rocchio, G (SC, past); Yepes, M (SC, past); Steiner, B (RC); Dulin, H (RC, past); Cayford, M (RS); Chu, K (RS); DeManbey, A (RS); Hillard, M (RS); Martin, K (RS); Thangthaeng, N (RS); Gurry, L (RS, past); Kochis, R (RS, past); Raymond, E (RS, past); Ripley, V (RS, past); Stevens, C (RS, past)

**MedStar Health Research Institute/ MedStar Baltimore:** Park, J (PI); Aroda, V (PI, past); Ghazi, A (Co-PI); Magee, M (Co-I); Ressing, Ann (Co-I); Loveland, A (SC); Hamm, M (SC); Hurtado, M (SC); Kuhn, A (SC); Leger, J (SC); Manandhar, L (SC); Mwicigi, F (SC); Sanchez, O (SC); Young, T (SC)

**Miami VA Healthcare System/University of Miami:** Garg, R (PI), Lagari-Libhaber, V (PI); Florez, HJ (PI, past); Valencia, WM (PI, past); Marks, J (Co-PI, past); Casula, S (Co-I); Oropesa-Gonzalez, L (SC); Hue, L (SC, past); Cuadot, A (SC, past); Nieto-Martinez, R (SC, past); Riccio Veliz, AK (SC, past); Gutt, M (RC, past); Kendal, YJ (RS, past); Veciana, B. (RS, past)

**Oregon Health & Science University:** Ahmann, A (PI); Aby-Daniel, D (Co-I); Joarder, F (Co-I); Morimoto, V (Co-I); Sprague, C (Co-I); Yamashita, D (Co-I); Cady, N (SC); Rivera-Eschright, N (SC); Kirchhoff, P (SC, past); Morales Gomez, B (RC); Adducci, J (RC, past); Goncharova, A (RC, past)

**Pacific Health Research and Education Institute/VA Pacific Islands:** Hox, SH (PI); Petrovitch, H (Co-PI); Matwichyna, M (SC); Jenkins, V (SC, past); Broadwater, L (RS); Ishii, RR (RS); Bermudez, NO (RS; past)

**Pennington Biomedical Research Center:** Hsia, DS (PI); Cefalu, WT (PI, past); Greenway, FL (Co-I); Waguespack, C (Co-I); King, E (SC); Fry, G (SC); Dragg, A (SC); Gildersleeve, B (SC); Arceneaux, J (SC); Haynes, N (SC, past); Thomassie, A (SC, past); Pavlionis, M (SC, past); Bourgeois, B (RC, past); Hazlett, C (RS)

**San Diego VA Medical Center:** Mudaliar, S (PI); Henry, R (PI, past); Boeder, S (Co-I, past); Pettus, J (Co-I, past); Diaz, E (SC); Garcia-Acosta, D (SC); Maggs, S (SC); DeLue, C (SC, past); Stallings, A (SC, past); Castro, E (RC, past); Hernandez, S (RC, past)

**Southwestern American Indian Center:** Krakoff, J (PI); Curtis, JM (Co-I); Killean, T (SC); Khalid, M (SC); Joshevama, E (RC, past); Diaz, E (RS); Martin, D (RS); Tsingine, K (RS); Karshner, T (RS, past)

**St. Luke's‐Roosevelt Hospital:** Albu, J (Co-PI); Pi-Sunyer, FX (Co-PI, past); Frances, S (Co-I); Maggio, C (SC, past); Ellis, E (RC); Bastawrose, J (RC, past); Gong, X (RS)

**SUNY Downstate Medical Center/New York Hospital‐Queens:** Banerji, MA (PI); August, P (Co-I); Lee, M (Co-I); Lorber, D (Co-I); Brown, NM (SC, RC); Josephson, DH (SC); Thomas, LL (SC, RC); Tsovian, M (SC, RC); Cherian, A (SC, RC, past); Jacobson, MH (RS); Mishko, MM (RS)

**The University of North Carolina Diabetes Care Center:** Kirkman, MS (PI); Buse, JB (Co-I); Diner, J (Co-I); Dostou, J (Co-I); Machineni, S (Co-I); Young, L (Co-I); Bergamo, K (Co-I, past); Goley, A (Co-I, past); Kerr, J (Co-I, past); Largay, JF (Co-I, past); Guarda, S (SC); Cuffee, J (SC, past); Culmer, D (SC, past); Fraser, R (RC); Almeida, H (RC, past); Coffer, S (RC, past); Debnam, E (RC, past); Kiker, L (RC, past); Morton, S (RC, past); Josey, K (RS); Fuller, G (RS, past)

**University of Alabama Birmingham:** Garvey, WT (PI); Cherrington, AL (Co-PI); Dyer, D (SC); Lawson, MCR (SC); Griffith, O (SC, past); Agne, A (RC); McCullars, S (RC)

**University of Cincinnati/Cincinnati VA Medical Center:** Cohen, RM (PI); Craig, J (SC); Rogge, MC (SC; past); Burton, K (SC, past); Kersey, K (SC, RC, past); Wilson, C (SC, past); Lipp, S (RC, past); Vonder Meulen, MB (RC, past); Adkins, C (RS); Onadeko, T (RS)

**University of Colorado‐Denver/VA:** Rasouli, N (PI); Baker, C (Co-I); Schroeder, E (Co-I, past); Razzaghi, M (Co-I); Lyon, C (Co-I, past); Penaloza, R (Co-I, past); Underkofler, C (SC); Lorch, R (SC); Douglass, S (SC, past); Steiner, S (SC, past)

**University of Iowa:** Sivitz, WI (PI); Cline, E (SC); Knosp, LK (SC); McConnell, J (SC, past); Lowe, T (RC)

**University of Michigan:** Herman, WH (PI); Pop-Busui, R (Co-PI); Tan, MH (Co-I); Martin, C (SC); Waltje, A (SC, RC); Katona, A (SC); Goodhall, L (SC, past); Eggleston, R (RC, past); Kuo, S (RS); Bojescu, S (RS, past); Bule, S (RS, past); Kessler, N (RS, past); LaSalle, E (RS, past); Whitley, K (RS, past)

**University of Minnesota:** Seaquist, ER (PI); Bantle, A (Co-I); Harindhanavudhi, T (Co-I); Kumar, A (Co-I); Redmon, B (Co-I); Bantle, J (Co-I, past); Coe, M (SC); Mech, M (SC); Taddese, A (RC); Lesne, L (RS); Smith, S (RS)

**University of Nebraska Medical Center/Omaha VA:** Desouza, C (PI); Kuechenmeister, L (Co-I); Shivaswamy, V (Co-I); Burbach, S (SC); Rodriguez, MG (SC); Seipel, K (SC); Alfred, A (SC, past); Morales, AL (SC; past); Eggert, J (RS); Lord, G (RS); Taylor, W (RS, past); Tillson, R (RS, past)

**University of New Mexico:** Schade, DS (PI); Adolphe, A (Co-PI); Burge, M (Co-PI, past); Duran-Valdez, E (SC); Martinez, J (RC, past); Bancroft, A (RS); Kunkel, S (RS); Ali Jamaleddin Ahmad, F (RS, past); Hernandez McGinnis, D (RS, past); Pucchetti, B (RS, past); Scripsick, E (RS, past); Zamorano, A (RS, past)

**UT Health San Antonio:** DeFronzo, RA (PI); Cersosimo, E (Co-PI); Abdul-Ghani, M (Co-I); Triplitt, C (Co-I); Juarez, D (SC); Garza, RI (SC, past); Verastiqui, H (SC, past); Wright, K (RC, past); Puckett, C (RS)

**University of Texas‐Southwestern Medical Center:** Raskin, P (PI); Rhee, C (Co-I, past); Abraham, S (SC); Jordan, LF (SC); Sao, S (SC); Morton, L (SC, past); Smith, O (SC, past); Osornio Walker, L (RC, past); Schnurr-Breen, L (RC, past); Ayala, R (RS); Kreymer, RB (RS); Sturgess, D (RS, past)

**VA Puget Sound Health Care System/University of Washington:** Utzschneider, KM (PI); Kahn, SE (Co-I); Alarcon-Casas Wright, L (Co-I); Boyko, EJ (Co-I); Tsai, EC (Co-I); Trence, DL (Co-I, past); Trikudanathan, S (Co-I, past); Fattaleh, BN (SC); Montgomery, BK (SC, past); Atkinson, KM (RS); Kozedub, A (RS); Concepcion, T (RS, past); Moak, C (RS, past); Prikhodko, N (RS, past); Rhothisen, S (RS, past)

**Vanderbilt University:** Elasy, TA (PI); Martin, S (SC); Shackelford, L (RC, RS, past); Goidel, R (RS); Hinkle, N (RS); Lovell, C (RS); Myers, J (RS); Lipps Hogan, J (RS, past)

**Washington University:** McGill, JB (PI); Salam, M (Co-I); Schweiger, T (SC, RC); Kissel, S (SC, RC, past); Recklein, C (SC, past); Clifton, MJ (RS)

**Yale University/Fair Haven Community Health Center/West Haven VA Medical Center:** Tamborlane, W (PI); Camp, A (Co-I); Gulanski, B (Co-I); Inzucchi, SE (Co-I); Pham, K (Co-I); Alguard, M (SC, RC); Gatcomb, P (SC); Lessard, K (SC); Perez, M (SC); Iannone, L (RC); Magenheimer, E (RC); Montosa, A (RC)

**Study Units**

**NIH/NIDDK (Sponsor):** Cefalu, WT (Director, Division of Diabetes, Endocrinology and Metabolic Diseases); Fradkin, J (Director, Division of Diabetes, Endocrinology and Metabolic Diseases, past); Burch, HB (Project Scientist); Bremer, AA (Project Scientist, past)

**Chairman’s Office, Massachusetts General Hospital, Harvard Medical School:** Nathan, DM (Study Chair, Study Co-PI)

**Executive Committee:** Nathan, DM (Study Chair, Study Co-PI); Lachin, JM (U01 Contact PI, Study Co-PI); Buse, JB (Co-I); Kahn, SE (Co-I); Krause-Steinrauf, H (Co-I, Project Director); Larkin, ME (Co-I, SC); Tiktin, M (Co-I, SC); Wexler, D (PI); Burch, HB (Program Scientist); Bremer, AA (Project Scientist, past)

**Coordinating Center, The George Washington University Biostatistics Center:** Lachin, JM (U01 Contact PI, Study Co-PI); Krause-Steinrauf, H (Co-I, Project Director); Younes, N (Co-I); Bebu, I (RS); Butera, N (RS); Buys, CJ (RS); Fagan, A (RS); Gao, Y (RS); Ghosh, A (RS); Gramzinski, MR (RS); Hall, SD (RS); Kazemi, E (RS); Legowski, E (RS); Liu, H (RS); Suratt, C (RS); Tripputi, M (RS); Arey, A (RS, past); Backman, M (RS, past); Bethepu, J (RS, past); Lund, C (RS, past); Mangat Dhaliwal, P (RS, past); McGee, P (RS, past); Mesimer, E (RS, past); Ngo, L (RS, past)

**Central Biochemical Laboratory, University of Minnesota Advanced Research and Diagnostic Laboratory:** Steffes, M (PI); Seegmiller, J (Co-I); Saenger, A (Co-I, past); Arends, V (SC); Gabrielson, D (SC, past)

**Drug Distribution Center, VA Cooperative Studies Program Clinical Research Pharmacy Coordinating Center:** Conner, T (PI); Warren, S (PI, past); Day, J (RS); Huminik, J (RS); Scrymgeour, A (RS)

**ECG Reading Center, EPICARE, Wake Forest University:** Soliman, EZ (PI); Pokharel, Y (PI, past), Zhang, ZM (Co-I, past); Campbell, C (SC); Hu, J (SC); Keasler, L (SC); Hensley, S (SC, past); Li, Y (RS)

**Economic Evaluation and Assessment Center:**

**University of Michigan:**  Herman, WH (PI); Kuo, S (RS); Martin, C (SC); Waltje, A (SC, RC); Mihalcea, R (RS); Min, DJ (RS); Perez-Rosas, V (RS); Prosser, L (RS); Resnicow, K (RS); Ye, W (RS)

**Centers for Disease Control and Prevention:** Shao, H (RS); Zhang, P (RS)

**Neurocognitive Coordinating Center, Columbia University Medical Center:** Luchsinger, J (PI); Sanchez, D (SC); Assuras, S (RS)

**QWB Reading Center, University of California San Diego Health Services Research Center:** Groessl, E (PI); Sakha, F (SC); Chong, H (SC, past); Hillery, N (RS)

**Collaborators**

**Cardiovascular Adjudication Advisor:** Everett, BM (Brigham and Women’s Hospital)

**Collaborating Investigators (Recruitment Sites):** Abdouch, I (University of Nebraska Medical Center/Omaha VA); Bahtiyar, G (SUNY Downstate Medical Center); Brantley, P (Pennington Biomedical Research Center (LSU)); Broyles, FE (Swedish Medical Center); Canaris, G (University of Nebraska Medical Center/Omaha VA); Copeland, P (Massachusetts General Hospital); Craine, JJ (UW Valley Medical Center); Fein, WL (Swedish Medical Center); Gliwa, A (SUNY Downstate Medical Center); Hope, L (SUNY Downstate Medical Center); Lee, MS (SUNY Downstate Medical Center); Meiners, R (Pennington Biomedical Research Center (LSU)); Meiners, V (Pennington Biomedical Research Center (LSU)); O’Neal, H (Pennington Biomedical Research Center (LSU)); Park, JE (UW Valley Medical Center); Sacerdote, A (SUNY Downstate Medical Center; Sledge, Jr., E (Pennington Biomedical Research Center (LSU)); Soni, L (SUNY Downstate Medical Center); Steppel-Reznik, J (Massachusetts General Hospital); Turchin, A (Massachusetts General Hospital)

**Beta Cell Ancillary Study:** Brooks-Worrell, B (University of Washington); Hampe, CS (University of Washington); Palmer, JP (University of Washington); Shojaie, A (University of Washington)

**Continuous Glucose Monitoring Sub-study:** Higgins, J (Massachusetts General Hospital; Harvard Medical School)

**Emotional Distress Sub-study:** Golden, S (Johns Hopkins University); Gonzalez, J (Yeshiva University; Albert Einstein College of Medicine); Naik, A (Baylor College of Medicine); Walker, E (Albert Einstein College of Medicine)

**National Diabetes Education Program (NDEP) Sub-study:** Doner Lotenberg, L (Hager Sharp); Gallivan, JM (National Institutes of Health); Lim, J (Hager Sharp); Tuncer, DM (National Institutes of Health)

**Recruitment Ancillary Study:** Behringer-Massera, S (The Mount Sinai Hospital, Beth Israel Medical Center)

### SUPPLEMENTARY TABLES

#### **Table S1.** Baseline characteristics of genotyped individuals in GRADE by treatment group.

|  | **Glargine** | **Glimepiride** | **Liraglutide** | **Sitagliptin** | ***p*-value** |
| --- | --- | --- | --- | --- | --- |
| ***Demographics*** |  |  |  |  |  |
| N | 1,131 | 1,154 | 1,134 | 1,153 |  |
| Age at baseline visit (years) | 57.3 ± 9.8 | 57.3 ± 10.0 | 57.7 ± 9.7 | 57.7 ± 9.9 | 0.551 |
| Age group (years) |  |  |  |  | 0.178 |
| <45 years | 126 (11.1%) | 147 (12.7%) | 129 (11.4%) | 120 (10.4%) |  |
| 45-59 years | 543 (48.0%) | 502 (43.5%) | 499 (44.0%) | 545 (47.3%) |  |
| 60+ years | 462 (40.8%) | 505 (43.8%) | 506 (44.6%) | 488 (42.3%) |  |
| Female sex | 389 (34.4%) | 432 (37.4%) | 382 (33.7%) | 410 (35.6%) | 0.258 |
| Self-reported Race |  |  |  |  | 0.111 |
| All others | 135 (11.9%) | 144 (12.5%) | 159 (14.0%) | 152 (13.2%) |  |
| Black or African American | 205 (18.1%) | 239 (20.7%) | 212 (18.7%) | 187 (16.2%) |  |
| White | 791 (69.9%) | 771 (66.8%) | 763 (67.3%) | 814 (70.6%) |  |
| Self-reported Ethnicity |  |  |  |  | 0.824 |
| Non-Hispanic | 920 (82.1%) | 923 (81.0%) | 911 (80.8%) | 924 (80.7%) |  |
| Hispanic | 201 (17.9%) | 217 (19.0%) | 217 (19.2%) | 221 (19.3%) |  |
| Education completed |  |  |  |  | 0.136 |
| Less than high school | 80 (7.1%) | 89 (7.7%) | 77 (6.8%) | 75 (6.5%) |  |
| High school/GED | 222 (19.6%) | 233 (20.2%) | 244 (21.5%) | 250 (21.7%) |  |
| Some college | 353 (31.2%) | 330 (28.6%) | 330 (29.1%) | 303 (26.3%) |  |
| College | 298 (26.3%) | 295 (25.6%) | 270 (23.8%) | 333 (28.9%) |  |
| Graduate School | 178 (15.7%) | 207 (17.9%) | 213 (18.8%) | 191 (16.6%) |  |
| ***Anthropometrics*** |  |  |  |  |  |
| Weight (kg) | 100.9 ± 22.5 | 99.9 ± 22.6 | 100.5 ± 23.0 | 99.7 ± 21.7 | 0.586 |
| Waist circumference (cm) | 112.8 ± 15.8 | 112.5 ± 16.1 | 112.8 ± 16.3 | 112.0 ± 15.0 | 0.594 |
| ***Blood pressure (BP)*** |  |  |  |  |  |
| Systolic (mmHg) | 128.5 ± 14.9 | 128.4 ± 14.5 | 128.3 ± 15.0 | 128.4 ± 14.9 | 0.995 |
| Diastolic (mmHg) | 77.4 ± 10.0 | 77.0 ± 9.6 | 77.3 ± 9.9 | 77.3 ± 9.7 | 0.797 |
| History of hypertension | 757 (66.9%) | 770 (66.7%) | 765 (67.5%) | 755 (65.5%) | 0.780 |
| ***Cardiovascular*** |  |  |  |  |  |
| History of MI | 62 (5.5%) | 54 (4.7%) | 59 (5.2%) | 68 (5.9%) | 0.616 |
| History of stroke | 27 (2.4%) | 27 (2.3%) | 18 (1.6%) | 25 (2.2%) | 0.528 |
| ***Diabetes*** |  |  |  |  |  |
| BMI (kg/m^2^) | 34.5 ± 6.9 | 34.3 ± 6.9 | 34.3 ± 6.8 | 34.1 ± 6.7 | 0.718 |
| Duration of diabetes  (years) | 4.0 ± 2.7 | 4.2 ± 2.8 | 4.1 ± 2.7 | 4.0 ± 2.7 | 0.336 |
| HbA1c (%) | 7.5 ± 0.5 | 7.5 ± 0.5 | 7.5 ± 0.5 | 7.5 ± 0.5 | 0.693 |
| Fasting glucose (mg/dL) | 152.9 ± 31.6 | 151.8 ± 31.4 | 151.1 ±30.5 | 151.1 ± 28.9 | 0.466 |
| Fasting insulin (mU/L) | 23.2 ± 15.2 | 21.8± 15.0 | 21.1 ± 14.0 | 21.8 ± 15.6 | 0.013 |
| ***Lipids*** |  |  |  |  |  |
| Total Cholesterol (mg/dL) | 163.7 ± 38.6 | 162.5 ± 37.2 | 164.3 ± 38.3 | 162.1 ± 37.2 | 0.468 |
| HDL (mg/dL) | 43.2 ± 10.5 | 43.4 ± 10.1 | 43.3 ± 11.0 | 43.2 ± 10.5 | 0.975 |
| LDL (mg/dL) | 91.1 ± 32.4 | 89.4 ± 31.3 | 90.7 ± 32.0 | 88.5 ± 30.8 | 0.193 |
| Triglycerides (mg/dL) | 150.8 ± 108.7 | 154.1 ± 126.6 | 158.9 ± 135.1 | 155.2 ± 116.3 | 0.471 |
| ***Renal*** |  |  |  |  |  |
| eGFR < 60 | 34 (3.0%) | 25 (2.2%) | 30 (2.6%) | 28 (2.4%) | 0.630 |
| Moderately elevated   albuminuria | 154 (13.6%) | 160 (13.9%) | 176 (15.5%) | 159 (13.8%) | 0.525 |
| Severely elevated   albuminuria | 16 (1.4%) | 25 (2.2%) | 19 (1.7%) | 14 (1.2%) | 0.298 |

Continuous outcomes were compared using t-tests, and categorical outcomes were analyzed using chi-square tests.

#### **Table S2.** Comparison of baseline characteristics between individuals included in genetic analyses to the remainder of the GRADE participants.

|  | **Did not consent to genetic analyses** | **Poor DNA quality or failed QC** | **Included in genetic analyses** | ***p*-value*** | ***p*-value^†^** |
| --- | --- | --- | --- | --- | --- |
| ***Demographics*** |  |  |  |  |  |
| N | 298 | 177 | 4,572 |  |  |
| Treatment Group |  |  |  | 0.287 | 0.220 |
| Glargine | 77 (25.8%) | 55 (31.1%) | 1,131 (24.7%) |  |  |
| Glimepiride | 61 (20.5%) | 39 (22.0%) | 1,154 (25.2%) |  |  |
| Liraglutide | 83 (27.9%) | 45 (25.4%) | 1,134 (24.8%) |  |  |
| Sitagliptin | 77 (25.8%) | 38 (21.5%) | 1,153 (25.2%) |  |  |
| Age at baseline visit (years) | 55.1 ± 9.8 | 51.9 ± 11.0 | 57.5 ± 9.9 | <0.001 | <0.001 |
| Age category (years) |  |  |  | <0.001 | <0.001 |
| <45 years | 44 (14.8%) | 53 (29.9%) | 522 (11.4%) |  |  |
| 45-59 years | 160 (53.7%) | 76 (42.9%) | 2,089 (45.7%) |  |  |
| 60+ years | 94 (31.5%) | 48 (27.1%) | 1,961 (42.9%) |  |  |
| Female sex | 136 (45.6%) | 88 (49.7%) | 1,613 (35.3%) | <0.001 | <0.001 |
| Self-reported Race |  |  |  | <0.001 | <0.001 |
| All others | 56 (18.8%) | 87 (49.2%) | 590 (12.9%) |  |  |
| Black or African American | 117 (39.3%) | 40 (22.6%) | 843 (18.4%) |  |  |
| White | 125 (41.9%) | 50 (28.2%) | 3,139 (68.7%) |  |  |
| Self-reported Ethnicity |  |  |  | 0.038 | 0.893 |
| Non-Hispanic | 255 (86.1%) | 144 (81.8%) | 3,678 (81.1%) |  |  |
| Hispanic | 41 (13.9%) | 32 (18.2%) | 856 (18.9%) |  |  |
| Education completed |  |  |  | 0.441 | 0.002 |
| Less than high school | 20 (6.7%) | 23 (13.0%) | 321 (7.0%) |  |  |
| High school/GED | 57 (19.1%) | 33 (18.6%) | 949 (20.8%) |  |  |
| Some college | 87 (29.2%) | 60 (33.9%) | 1,316 (28.8%) |  |  |
| College | 91 (30.5%) | 45 (25.4%) | 1,196 (26.2%) |  |  |
| Graduate School | 43 (14.4%) | 16 (9.0%) | 789 (17.3%) |  |  |
| ***Anthropometrics*** |  |  |  |  |  |
| Weight (kg) | 98.1 ± 20.0 | 95.2 ± 21.7 | 100.3 ± 22.5 | 0.077 | 0.003 |
| Waist circumference (cm) | 110.5 ± 15.4 | 110.3 ± 15.3 | 112.5 ± 15.8 | 0.025 | 0.054 |
| ***Blood pressure (BP)*** |  |  |  |  |  |
| Systolic (mmHg) | 128.4 ± 14.6 | 126.5 ± 13.4 | 128.4 ± 14.8 | 0.993 | 0.060 |
| Diastolic (mmHg) | 77.8 ± 10.1 | 77.8 ± 10.2 | 77.2 ± 9.8 | 0.329 | 0.457 |
| History of hypertension | 199 (66.8%) | 93 (52.5%) | 3,047 (66.6%) | 1.000 | <0.001 |
| ***Cardiovascular*** |  |  |  |  |  |
| History of MI | 9 (3.0%) | 3 (1.7%) | 243 (5.3%) | 0.110 | 0.050 |
| History of stroke | 4 (1.3%) | 0 (0.0%) | 97 (2.1%) | 0.481 | 0.092 |
| ***Diabetes*** |  |  |  |  |  |
| BMI (kg/m^2^) | 34.0 ± 6.7 | 34.0 ± 6.7 | 34.3 ± 6.8 | 0.368 | 0.589 |
| Duration of diabetes  (years) | 3.9 ± 2.8 | 3.7 ± 2.8 | 4.1 ± 2.7 | 0.306 | 0.068 |
| HbA1c (%) | 7.5 ± 0.5 | 7.6 ± 0.5 | 7.5 ± 0.5 | 0.894 | 0.002 |
| Fasting glucose (mg/dL) | 148.6 ± 30.6 | 150.7 ± 37.3 | 151.7 ± 30.6 | 0.094 | 0.709 |
| Fasting insulin (mU/L) | 21.2 ± 16.7 | 21.0 ± 13.0 | 22.0 ± 15.0 | 0.506 | 0.385 |
| ***Lipids*** |  |  |  |  |  |
| Total Cholesterol (mg/dL) | 169.7 ± 35.8 | 170.6 ± 37.8 | 163.2 ± 37.8 | 0.003 | 0.011 |
| HDL (mg/dL) | 44.6 ± 11.1 | 45.1 ± 10.4 | 43.3 ± 10.5 | 0.046 | 0.026 |
| LDL (mg/dL) | 97.2 ± 32.2 | 95.0 ± 31.4 | 89.9 ± 31.6 | <0.001 | 0.039 |
| Triglycerides (mg/dL) | 140.2 ± 94.0 | 157.7 ± 146.8 | 154.7 ± 122.0 | 0.012 | 0.795 |
| ***Renal*** |  |  |  |  |  |
| eGFR < 60 | 4 (1.3%) | 4 (2.3%) | 117 (2.6%) | 0.265 | 0.996 |
| Moderately elevated   albuminuria | 41 (13.9%) | 26 (14.7%) | 649 (14.2%) | 0.932 | 0.945 |
| Severely elevated   albuminuria | 7 (2.4%) | 3 (1.7%) | 74 (1.6%) | 0.462 | 1.000 |
| ***Clinical Site*^‡^** |  |  |  | <0.001 | <0.001 |
| A | 5 (3.2%) | 0 (0.0%) | 149 (96.8%) |  |  |
| B | 0 (0.0%) | 0 (0.0%) | 28 (100.0%) |  |  |
| C | 1 (1.2%) | 0 (0.0%) | 81 (98.8%) |  |  |
| D | 6 (5.9%) | 1 (1.0%) | 94 (93.1%) |  |  |
| E | 1 (0.9%) | 2 (1.8%) | 110 (97.3%) |  |  |
| F | 2 (4.2%) | 2 (4.2%) | 44 (91.7%) |  |  |
| G | 23 (16.5%) | 8 (5.8%) | 108 (77.7%) |  |  |
| H | 6 (5.2%) | 6 (5.2%) | 103 (89.6%) |  |  |
| I | 4 (2.6%) | 3 (2.0%) | 146 (95.4%) |  |  |
| J | 18 (16.5%) | 2 (1.8%) | 89 (81.7%) |  |  |
| K | 5 (5.3%) | 1 (1.1%) | 88 (93.6%) |  |  |
| L | 3 (2.7%) | 1 (0.9%) | 106 (96.4%) |  |  |
| M | 6 (7.5%) | 2 (2.5%) | 72 (90.0%) |  |  |
| N | 5 (3.9%) | 4 (3.1%) | 119 (93.0%) |  |  |
| O | 8 (4.8%) | 8 (4.8%) | 152 (90.5%) |  |  |
| P | 2 (1.7%) | 1 (0.8%) | 118 (97.5%) |  |  |
| Q | 1 (0.6%) | 5 (3.2%) | 151 (96.2%) |  |  |
| R | 12 (8.6%) | 2 (1.4%) | 126 (90.0%) |  |  |
| S | 0 (0.0%) | 2 (1.6%) | 123 (98.4%) |  |  |
| T | 26 (17.9%) | 42 (29.0%) | 77 (53.1%) |  |  |
| U | 2 (5.0%) | 3 (7.5%) | 35 (87.5%) |  |  |
| V | 3 (3.2%) | 9 (9.5%) | 83 (87.4%) |  |  |
| W | 2 (5.6%) | 3 (8.3%) | 31 (86.1%) |  |  |
| X | 0 (0.0%) | 0 (0.0%) | 11 (100.0%) |  |  |
| Y | 2 (1.1%) | 3 (1.6%) | 183 (97.3%) |  |  |
| Z | 5 (9.8%) | 2 (3.9%) | 44 (86.3%) |  |  |
| AA | 3 (5.7%) | 0 (0.0%) | 50 (94.3%) |  |  |
| AB | 4 (3.0%) | 0 (0.0%) | 128 (97.0%) |  |  |
| AC | 2 (1.9%) | 2 (1.9%) | 102 (96.2%) |  |  |
| AD | 0 (0.0%) | 1 (5.3%) | 18 (94.7%) |  |  |
| AE | 26 (13.8%) | 7 (3.7%) | 156 (82.5%) |  |  |
| AF | 10 (6.8%) | 3 (2.0%) | 134 (91.2%) |  |  |
| AG | 12 (15.8%) | 1 (1.3%) | 63 (82.9%) |  |  |
| AH | 4 (6.6%) | 1 (1.6%) | 56 (91.8%) |  |  |
| AI | 8 (6.6%) | 12 (9.9%) | 101 (83.5%) |  |  |
| AJ | 1 (0.7%) | 2 (1.3%) | 149 (98.0%) |  |  |
| AK | 0 (0.0%) | 11 (14.3%) | 66 (85.7%) |  |  |
| AL | 7 (6.1%) | 7 (6.1%) | 101 (87.8%) |  |  |
| AM | 14 (9.0%) | 4 (2.6%) | 138 (88.5%) |  |  |
| AN | 7 (4.0%) | 3 (1.7%) | 165 (94.3%) |  |  |
| AO | 3 (3.2%) | 1 (1.1%) | 89 (95.7%) |  |  |
| AP | 28 (13.4%) | 2 (1.0%) | 179 (85.6%) |  |  |
| AQ | 6 (4.0%) | 0 (0.0%) | 145 (96.0%) |  |  |
| AR | 2 (2.2%) | 0 (0.0%) | 88 (97.8%) |  |  |
| AS | 13 (6.7%) | 8 (4.1%) | 173 (89.2%) |  |  |

Continuous outcomes were compared using t-tests, and categorical outcomes were analyzed using chi-square tests. **^*^***p*-value compares baseline characteristics between individuals included in genetic analyses to those who did not consent to genetic analyses. **^†^***p*-value compares baseline characteristics between individuals included in genetic analyses to those who had poor DNA quality or failed quality control (QC). **^‡^**Clinical sites are labeled with alphabetical codes to ensure de-identification and preserve confidentiality.

#### **Table S3.** Replication of genome-wide significant variants associated with glimepiride response in published SU GWAS.

| **rsID** | **Chr** | **Position^*^** | **NEA** | **EA** | **EAF^†^** | **GRADE results** | | | | | **SU GWAS results** | |
| --- | --- | --- | --- | --- | --- | --- | --- | --- | --- | --- | --- | --- |
|  |  |  |  |  |  |  | **Primary outcome** | | **Secondary outcome** | | **1-year change in HbA1c** | |
|  |  |  |  |  |  | **N** | **HR^‡^**  **(95% CI)** | ***p*-value** | **HR^‡^**  **(95% CI)** | ***p*-value** | **Beta**^§^  **(SE)** | ***p*-value** |
| rs1340183 | 1 | 178023490 | T | C | 0.839 | 1139 | 0.68  (0.6-0.78) | **1.02 × 10^-8^** | 0.76  (0.65-0.88) | 3.98 × 10^-4^ | 0.02 (0.03) | 0.54 |
| rs13001604 | 2 | 141017665 | G | A | 0.022 | 1139 | 2.41  (1.77-3.29) | **2.86 × 10^-8^** | 2.22  (1.57-3.15) | 6.67 × 10^-6^ | -0.08 (0.07) | 0.21 |
| rs115684396 | 2 | 19520878 | G | A | 0.022 | 1139 | 1.65  (1.19-2.3) | 3.05 × 10^-3^ | 2.70  (1.9-3.84) | **2.94 × 10^-8^** | 0.16 (0.10) | 0.11 |
| rs59676817 | 3 | 104825863 | G | A | 0.015 | 1139 | 3.35  (2.24-5.01) | **3.66 × 10^-9^** | 2.70  (1.72-4.23) | 1.57 × 10^-5^ | NA | NA |
| rs1905505 | 3 | 170977637 | G | A | 0.342 | 1139 | 1.36  (1.22-1.52) | **4.83 × 10^-8^** | 1.32  (1.16-1.51) | 2.52 × 10^-5^ | -0.04 (0.02) | **0.04** |
| rs11925227 | 3 | 171048829 | G | A | 0.170 | 1139 | 1.45  (1.28-1.65) | **4.87 × 10^-9^** | 1.30  (1.12-1.52) | 5.19 × 10^-4^ | -0.02 (0.03) | 0.43 |
| rs75718886 | 16 | 86575425 | C | G | 0.026 | 1139 | 2.41  (1.78-3.25) | **1.12 × 10^-8^** | 1.76  (1.23-2.5) | 1.74 × 10^-3^ | -0.25 (0.59) | 0.68 |
| rs62201522 | 20 | 38322086 | T | C | 0.030 | 1139 | 2.24  (1.69-2.98) | **2.43 × 10^-8^** | 1.55  (1.12-2.16) | 9.00 × 10^-3^ | 0.16 (0.33) | 0.63 |

NEA=Non-effect allele; EA=Effect allele; EAF=Effect allele frequency. NA indicates that the genetic variant was not available in the published sulfonylurea GWAS. **^*^**GRCh38 assembly. **^†^**This is the effect allele frequency based on the number of participants (N) in the GRADE results. ^‡^Hazard Ratios (HR) are expressed per 1 copy increase in the number of effect alleles (e.g. risk(1 EA)/risk(0 EA) or risk(2 EA)/risk(1EA))

^§^Beta represents the 1-year HbA1c reduction (baseline HbA1c minus follow-up HbA1c) per 1 copy increase in the number of effect alleles. A negative beta value indicates that the effect allele is associated with worse glycemic response.

#### **Table S4.** Replication of genome-wide significant variants associated with liraglutide response in published GLP-1 receptor agonist GWAS.

| **rsID** | **Chr** | **Position^*^** | **NEA** | **EA** | **EAF^†^** | **GRADE results** | | | | | **GLP-1 RA results** | |
| --- | --- | --- | --- | --- | --- | --- | --- | --- | --- | --- | --- | --- |
|  |  |  |  |  |  |  | **Primary outcome** | | **Secondary outcome** | | **1-year change in HbA1c** | |
|  |  |  |  |  |  | **N** | **HR**  **(95% CI)** | ***p*-value** | **HR**  **(95% CI)** | ***p*-value** | **Beta**  **(SE)** | ***p*-value** |
| rs80157733 | 3 | 122647534 | C | G | 0.048 | 1126 | 1.46  (1.16-1.83) | 1.05 × 10^-3^ | 2.09  (1.62-2.68) | **9.89 × 10^-9^** | -0.06 (0.04) | 0.24 |
| rs9934674 | 16 | 51378943 | T | C | 0.013 | 1126 | 2.37  (1.59-3.54) | 2.32 × 10^-5^ | 3.62  (2.32-5.65) | **1.45 × 10^-8^** | NA | NA |
| rs115566325 | 20 | 53780747 | C | T | 0.040 | 1126 | 2.15  (1.64-2.84) | **4.64 × 10^-8^** | 2.66  (1.94-3.66) | **1.66 × 10^-9^** | NA | NA |
| rs187004679 | 23 | 142526758 | T | C | 0.018 | 1126 | 2.60  (1.89-3.58) | **5.20 × 10^-9^** | 2.99  (2.12-4.22) | **4.52 × 10^-10^** | NA | NA |

NEA=Non-effect allele; EA=Effect allele; EAF=Effect allele frequency. NA indicates that the genetic variant was not available in the published GLP-1 receptor agonist GWAS. **^*^**GRCh38 assembly. ^†^This is the effect allele frequency based on the number of participants (N) in the GRADE results.

#### **Table S5.** Replication of genome-wide significant variants associated with glargine response in ORIGIN.

| **rsID** | **Chr** | **Position^*^** | **NEA** | **EA** | **EAF^†^** | **GRADE results** | | | | | **ORIGIN results** | | |
| --- | --- | --- | --- | --- | --- | --- | --- | --- | --- | --- | --- | --- | --- |
|  |  |  |  |  |  |  | **Primary outcome** | | **Secondary outcome** | | **1-year change in HbA1c** | | |
|  |  |  |  |  |  | **N** | **HR**  **(95% CI)** | ***p*-value** | **HR**  **(95% CI)** | ***p*-value** | **N** | **Beta**  **(SE)** | ***p*-value** |
| rs112620929 | 5 | 145825152 | G | A | 0.023 | 1125 | 1.49  (1.06-2.1) | 2.35 × 10^-2^ | 2.83  (1.96-4.10) | **3.30 × 10^-8^** | 1750 | NA | NA |
| rs73191136 | 7 | 108994863 | C | T | 0.025 | 1125 | 2.29  (1.71-3.08) | **3.53 × 10^-8^** | 2.10  (1.43-3.08) | 1.54 × 10^-4^ | 1750 | -0.23 (0.08) | **0.01** |
| rs76139219 | 8 | 119978123 | G | A | 0.028 | 1125 | 2.22  (1.7-2.91) | **6.89 × 10^-9^** | 1.65  (1.15-2.38) | 7.04 × 10^-3^ | 1750 | -0.22 (0.09) | **0.02** |
| rs75737981 | 19 | 38423936 | G | A | 0.018 | 1125 | 2.09  (1.46-3) | 5.74 × 10^-5^ | 3.17  (2.11-4.78) | **3.22 × 10^-8^** | 1750 | 0.17 (0.28) | 0.54 |
| rs10423332 | 19 | 41388347 | A | G | 0.015 | 1125 | 2.64  (1.79-3.9) | 1.09 × 10^-6^ | 3.50  (2.27-5.41) | **1.68 × 10^-8^** | 1750 | NA | NA |
| rs170675 | 21 | 20746463 | G | A | 0.030 | 1125 | 2.23  (1.67-2.96) | **4.24 × 10^-8^** | 1.62  (1.14-2.31) | 7.45 × 10^-3^ | 1750 | -0.10 (0.18) | 0.58 |

NEA=Non-effect allele; EA=Effect allele; EAF=Effect allele frequency. NA indicates that the genetic variant was not available in the ORIGIN genotyping data. ^*^GRCh38 assembly. ^†^This is the effect allele frequency based on the number of participants (N) in the GRADE results.

#### **Table S6.** Replication of genome-wide significant variants associated with sitagliptin response in SAVOR-TIMI 53.

| **rsID** | **Chr** | **Position^*^** | **NEA** | **EA** | **EAF^†^** | **GRADE results** | | | | | **SAVOR-TIMI 53 results** | | |
| --- | --- | --- | --- | --- | --- | --- | --- | --- | --- | --- | --- | --- | --- |
|  |  |  |  |  |  |  | **Primary outcome** | | **Secondary outcome** | | **1-year change in HbA1c** | | |
|  |  |  |  |  |  | **N** | **HR**  **(95% CI)** | ***p*-value** | **HR**  **(95% CI)** | ***p*-value** | **N** | **Beta**  **(SE)** | ***p*-value** |
| rs41265765 | 1 | 160123902 | G | T | 0.014 | 1147 | 2.37  (1.56-3.6) | 4.79 × 10^-5^ | 3.39  (2.2-5.21) | **2.87 × 10^-8^** | 3776 | 0.02 (0.09) | 0.82 |
| rs4963701 | 12 | 23552823 | A | G | 0.084 | 1147 | 1.17  (0.99-1.39) | 7.21 × 10^-2^ | 1.70  (1.4-2.05) | **4.60 × 10^-8^** | 3776 | 0.02 (0.04) | 0.54 |
| rs11409687 | 21 | 26800437 | C | CA | 0.028 | 1147 | 1.82  (1.38-2.4) | 2.54 × 10^-5^ | 2.40  (1.76-3.27) | **3.17 × 10^-8^** | 3776 | 0.02 (0.09) | 0.86 |

NEA=Non-effect allele; EA=Effect allele; EAF=Effect allele frequency. ^*^GRCh38 assembly. ^†^This is the effect allele frequency based on the number of participants (N) in the GRADE results.

#### **Table S7.** Conditional analyses for rs119255227 and rs1905505 in all participants and stratified by self-reported race.

| **Model** | **HR (95% CI)** | ***p*-value** |
| --- | --- | --- |
| *Conditioning on rs119255227* |  |  |
| rs1905505 (All) | 1.36 (1.22, 1.52) | **5×10^-8^** |
| rs1905505 (White) | 1.45 (1.27, 1.66) | **5×10^-8^** |
| rs1905505 (Black or African American) | 1.06 (0.86, 1.32) | 0.57 |
| rs1905505 \| rs11925227 (All) | 1.22 (1.06, 1.39) | **5×10^-3^** |
| rs1905505 \| rs11925227 (White) | 1.40 (1.16, 1.68) | **3×10^-4^** |
| rs1905505 \| rs11925227 (Black or African American) | 1.02 (0.80, 1.30) | 0.85 |
| *Conditioning on rs1905505* |  |  |
| rs11925227 (All) | 1.45 (1.28, 1.65) | **5×10^-9^** |
| rs11925227 (White) | 1.42 (1.22, 1.66) | **6×10^-6^** |
| rs11925227 (Black or African American) | 1.41 (1.08, 1.83) | **1×10^-2^** |
| rs11925227 \| rs1905505 (All) | 1.27 (1.09, 1.49) | **2×10^-3^** |
| rs11925227 \| rs1905505 (White) | 1.13 (0.92, 1.39) | 0.24 |
| rs11925227 \| rs1905505 (Black or African American) | 1.26 (0.93, 1.72) | 0.13 |

### SUPPLEMENTARY FIGURES

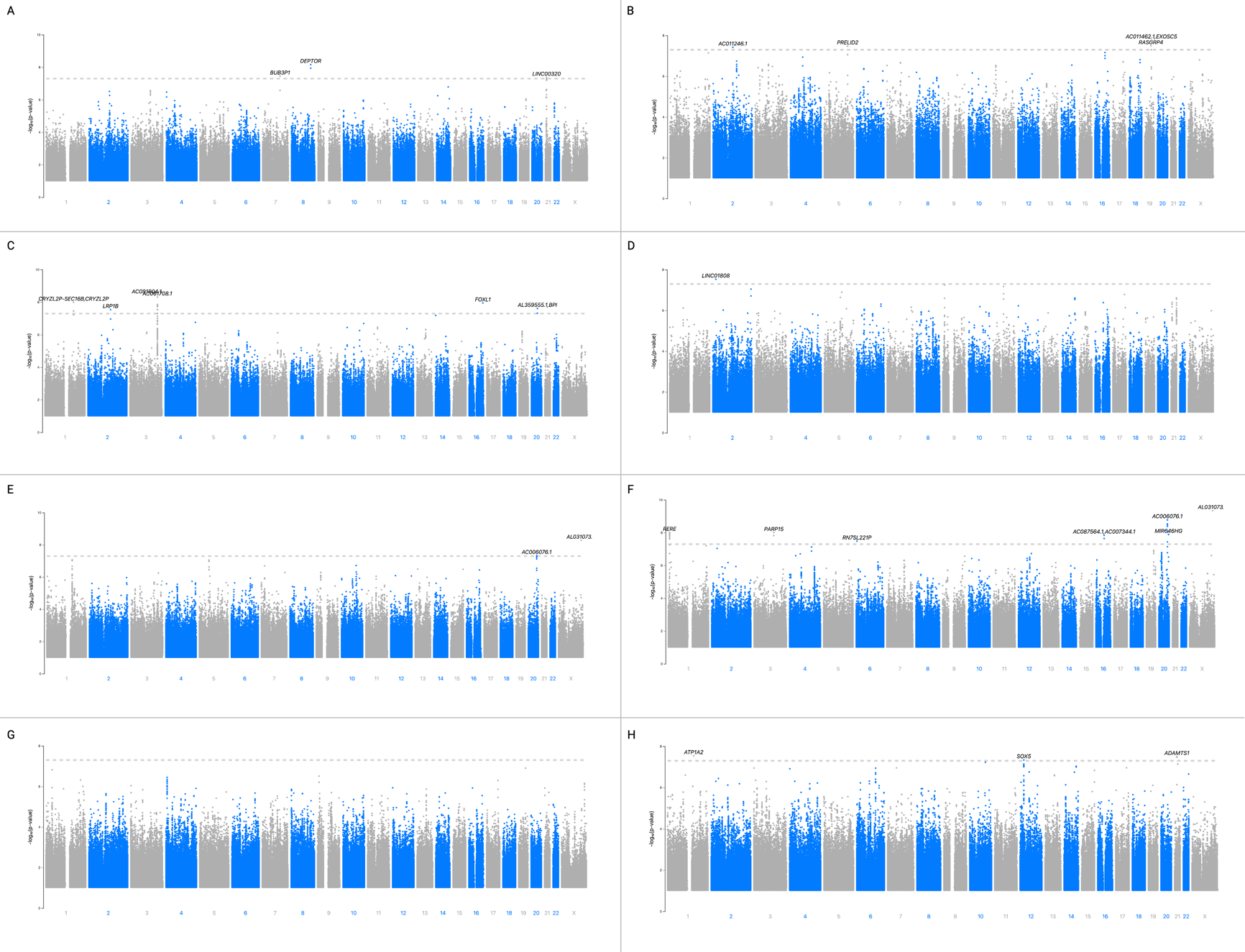

**Figure S1.** Manhattan plots for conducted GWAS analyses. A. Glargine – primary outcome; B. Glargine – secondary outcome; C. Glimepiride – primary outcome; D. Glimepiride – secondary outcome; E. Liraglutide – primary outcome; F. Liraglutide – secondary outcome; G. Sitagliptin – primary outcome; H. Sitagliptin – secondary outcome.

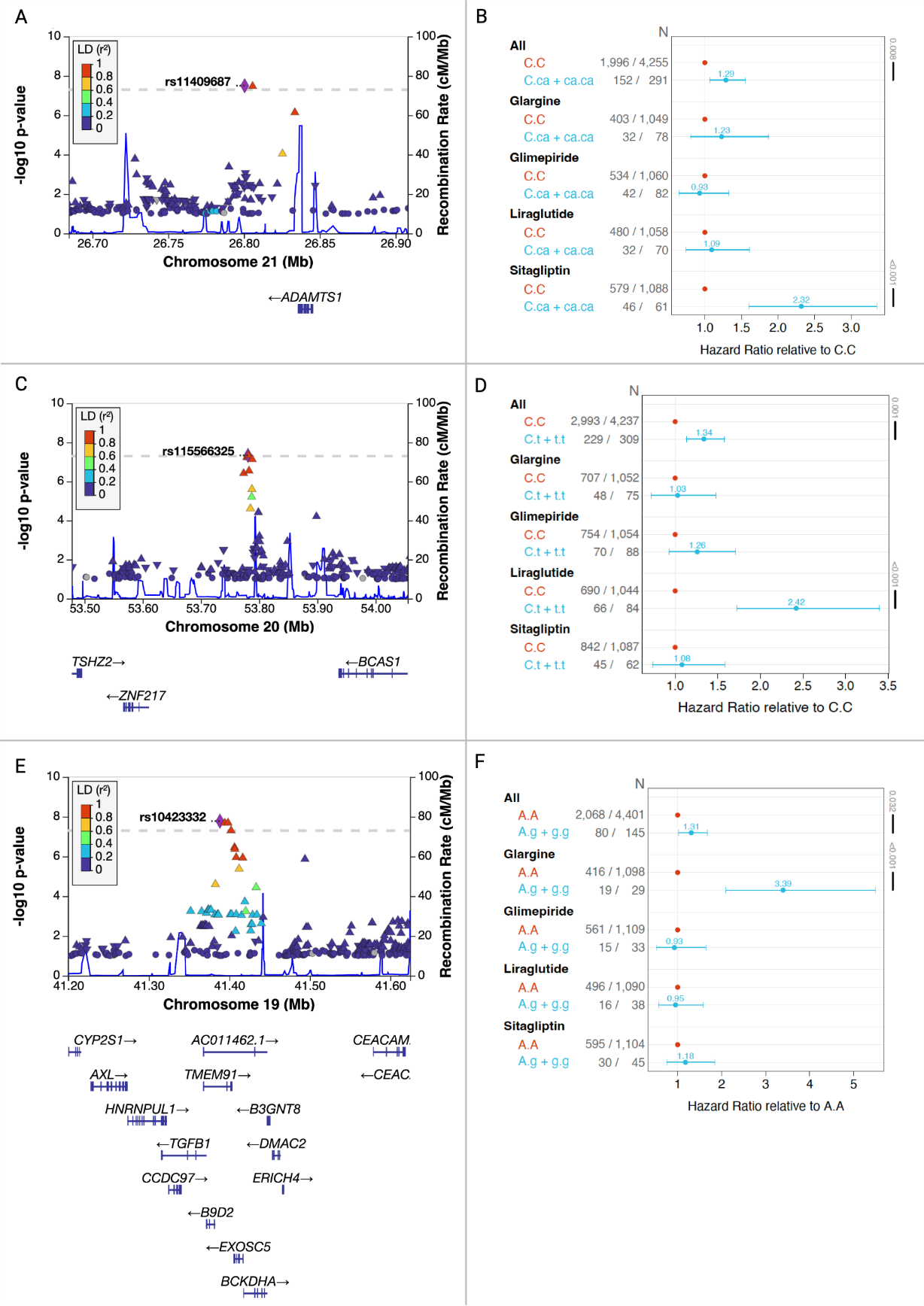

**Figure S2.** Selected genetic associations for treatment failure for sitagliptin, liraglutide, and glargine. A) Regional association plot of rs11409687 for secondary metabolic outcome in the sitagliptin arm. B) Hazard ratios (HRs) of treatment failure, as measured by reaching the secondary metabolic outcome, stratified by treatment arm and rs11409687 genotype. HRs are reported relative to non-carriers of the CA effect allele. C) Regional association plot of rs115566325 for primary metabolic outcome in the liraglutide arm. D) HRs of treatment failure, as measured by reaching the primary metabolic outcome, stratified by treatment arm and rs115566325 genotype. HRs are reported relative to non-carriers of the T effect allele. E) Regional association plot of rs10423332 for secondary metabolic outcome in the glargine arm. F) HRs of treatment failure, as measured by reaching the secondary metabolic outcome, stratified by treatment arm and rs10423332 genotype. HRs are reported relative to non-carriers of the G effect allele.

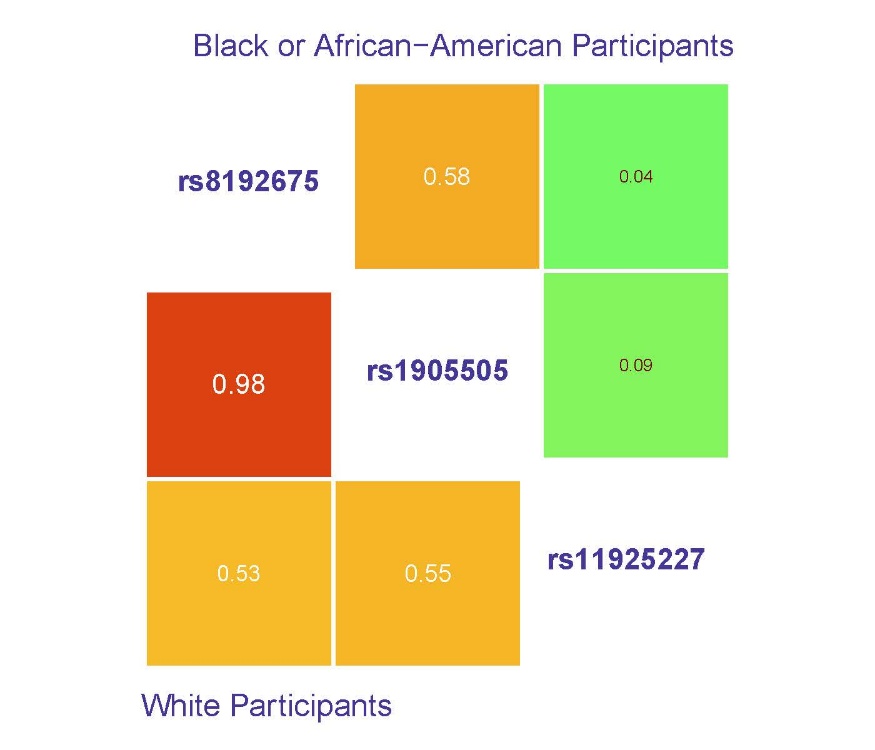

**Figure S3.** Linkage disequilibrium (R²) between rs1905505, rs11925227, and rs8192675 variants by self-reported race. R^2^ values are reported for each pair. Black or African-American participants are located in upper right, White participants in lower left.

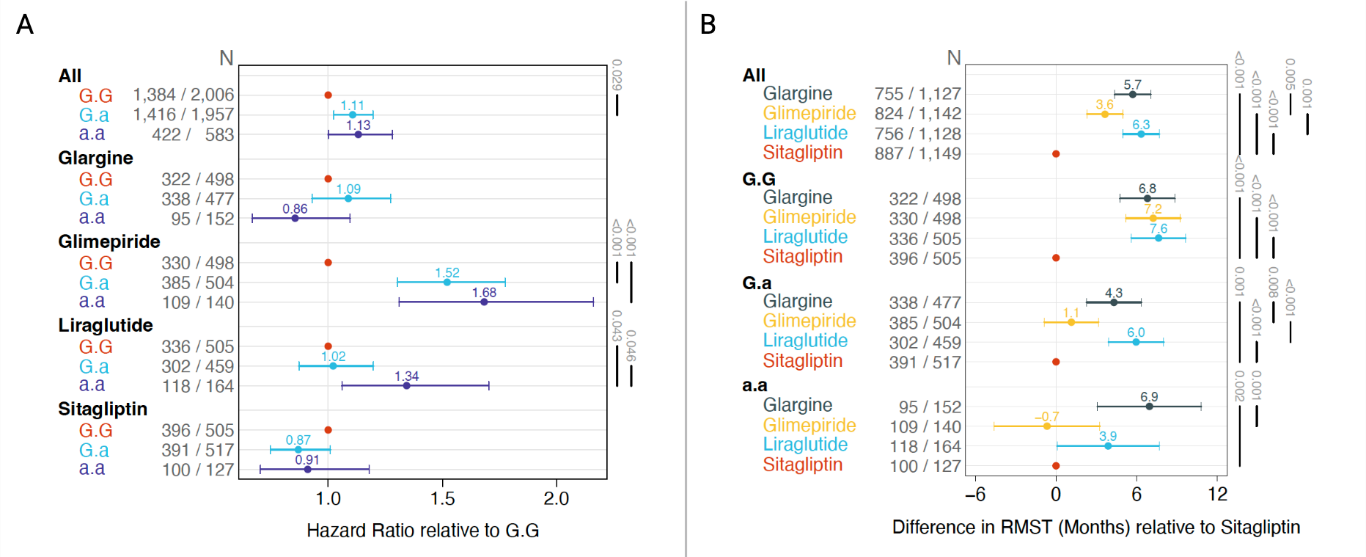

**Figure S4.** Treatment failure by rs1905505 genotype and treatment arm. A) Hazard ratios (HRs) of treatment failure, as measured by reaching the primary metabolic outcome, stratified first by treatment arm, followed by rs1905505 genotype. Non-carriers of the A effect allele is the reference group with a HR of 1.0. B) Restricted mean survival time (RMST) of treatment failure, as measured by months to reaching the primary metabolic outcome, stratified first by rs1905505 genotype, followed by treatment arm. Sitagliptin is the reference group with an RMST of 0.

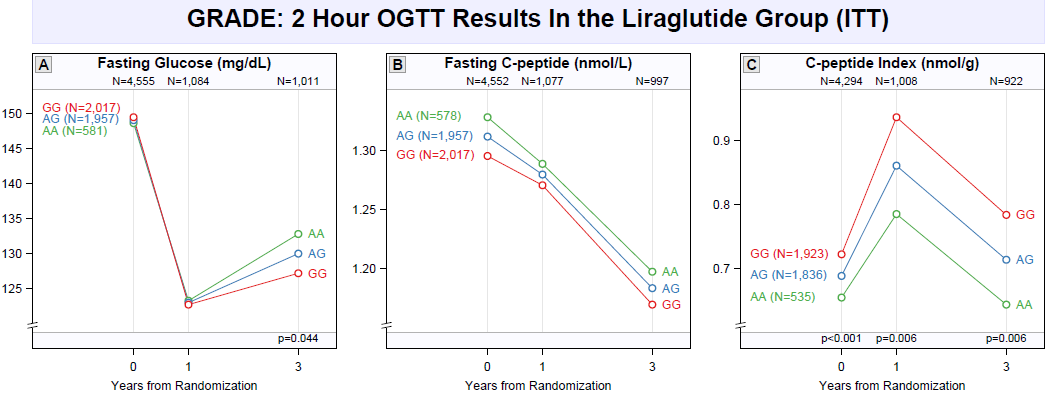

**Figure S5.** 2-hour OGTT results in the liraglutide group by intention-to-treat (ITT) analysis in GRADE. A) Fasting glucose, B) Fasting C-peptide, and C) C-peptide index (CPI) during the OGTT performed at years 0, 1, and 3 from randomization in GRADE by rs1905505 genotype. OGTT data at years 1 and 3 are displayed for the liraglutide arm based on ITT analysis. The reported N’s for AA, GG, and AA represents the distribution rs1905505 genotype at baseline prior to treatment assignment. *P*-values compare race-adjusted means of fasting glucose, fasting C-peptide, and CPI by rs1905505 genotype using an additive genetic model.

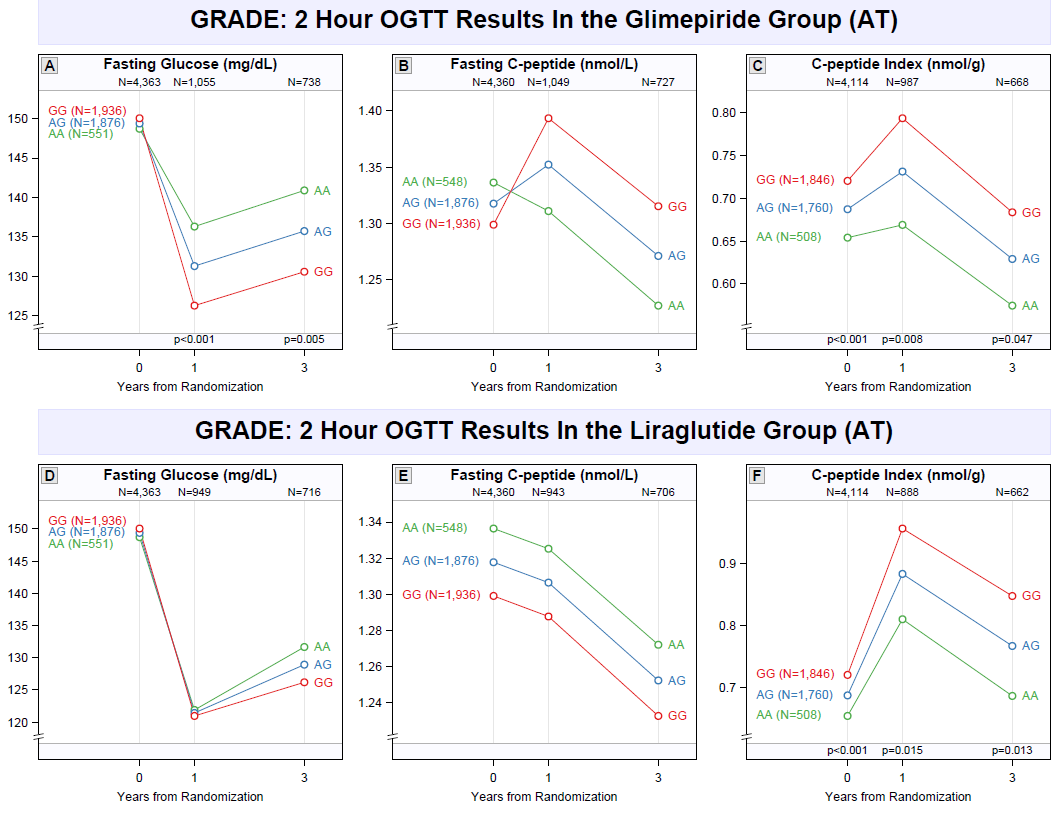

**Figure S6.** 2-hour OGTT results in glimepiride and liraglutide group and by as-treated (AT) analysis in GRADE. A) Fasting glucose, B) Fasting C-peptide, and C) C-peptide index (CPI) during the OGTT performed at years 0, 1, and 3 from randomization in GRADE by rs1905505 genotype. OGTT data at years 1 and 3 are displayed for the glimepiride arm based on AT analysis. D) Fasting glucose, E) Fasting C-peptide, and F) C-peptide index (CPI) during the OGTT performed at years 0, 1, and 3 from randomization in GRADE by rs1905505 genotype. OGTT data at years 1 and 3 are displayed for the liraglutide arm based on AT analysis. The reported N’s for AA, GG, and AA represents the distribution rs1905505 genotype at baseline prior to treatment assignment. *P*-values compare race-adjusted means of fasting glucose, fasting C-peptide, and CPI by rs1905505 genotype using an additive genetic model.

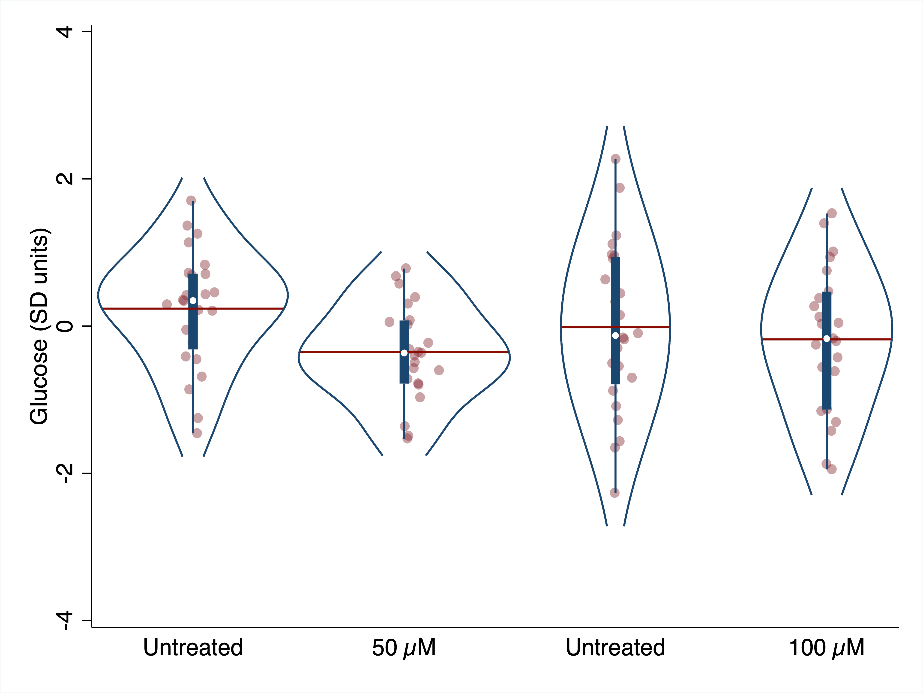

**Figure S7:** Effect of 50 and 100 µM glimepiride on transformed random glucose in zebrafish larvae. The violin plot shows the results of two pilot experiments examining the effect of 50 and 100 µM glimepiride on inverse normally transformed random glucose content in 24 larvae per group. Each concentration has its own controls because we reached maximum solubility of the drug in DMSO. This way we ensured that in each comparison, treated and untreated larvae were exposed to the same concentration of DMSO (0.818% for 50 µM glimepiride; 1.635% for 100 µM glimepiride).
